# Prediction models for post-discharge mortality among under-five children with suspected sepsis in Uganda: A multicohort analysis

**DOI:** 10.1101/2023.06.14.23291343

**Authors:** Matthew O Wiens, Vuong Nguyen, Jeffrey N Bone, Elias Kumbakumba, Stephen Businge, Abner Tagoola, Sheila Oyella Sherine, Emmanuel Byaruhanga, Edward Ssemwanga, Celestine Barigye, Jesca Nsungwa, Charles Olaro, J Mark Ansermino, Niranjan Kissoon, Joel Singer, Charles P Larson, Pascal M Lavoie, Dustin Dunsmuir, Peter P Moschovis, Stefanie Novakowski, Clare Komugisha, Mellon Tayebwa, Douglas Mwesigwa, Nicholas West, Martina Knappett, Nathan Kenya Mugisha, Jerome Kabakyenga

## Abstract

**Background:** In many low-income countries, more than five percent of hospitalized children die following hospital discharge. The identification of those at risk has limited progress to improve outcomes. We aimed to develop algorithms to predict post-discharge mortality among children admitted with suspected sepsis.

**Methods:** Four prospective cohort studies were conducted at six hospitals in Uganda between 2012 and 2021. Death occurring within six months of discharge was the primary outcome. Separate models were developed for children 0-6 months of age and for those 6-60 months of age, based on candidate predictors collected at admission. Within each age group, three models were derived, each with a maximum of eight variables based on variable importance. Deriving parsimonious models with different sets of predictors was prioritized to improve usability and support implementation in settings where some data elements are unavailable. All models were internally validated using 10-fold cross validation.

**Findings:** 8,810 children were prospectively enrolled, of whom 470 died in hospital and 161 (1·9%) were lost to follow-up; 257 (7·7%) and 233 (4·8%) post-discharge deaths occurred in the 0-6-month and 6-60-month age groups, respectively. The primary models had an area under the receiver operating characteristic curve (AUROC) of 0·77 (95%CI 0·74-0·80) for 0-6-month-olds and 0·75 (95%CI 0·72-0·79) for 6-60-month-olds; mean AUROCs among the 10 cross-validation folds were 0·75 and 0·73, respectively. Calibration across risk strata were good with Brier scores of 0·07 and 0·04, respectively. The most important variables included anthropometry and oxygen saturation. Additional variables included duration of illness, jaundice-age interaction, and a bulging fontanelle among 0-6-month-olds; and prior admissions, coma score, temperature, age-respiratory rate interaction, and HIV status among 6-60-month-olds.

**Interpretation:** Simple prediction models at admission with suspected sepsis can identify children at risk of post-discharge mortality. Further external validation is recommended for different contexts. Models can be integrated into existing processes to improve peri-discharge care as children transition from the hospital to the community.

**Funding:** Grand Challenges Canada (#TTS-1809-1939), Thrasher Research Fund (#13878), BC Children’s Hospital Foundation, and Mining4Life.

## Introduction

Morbidity and mortality secondary to sepsis disproportionately affect children, especially those in low- and middle-income countries, where over 85% of global cases and deaths occur.^1^ Lower income regions are plagued by poorly resilient health systems, widespread socio-economic deprivation and unique vulnerabilities such as malnutrition. In this context, reducing the overall burden of sepsis requires a multi-pronged strategy that addresses all three periods along the sepsis care continuum – pre-facility, facility and post-facility.^2^ Of these aspects, post-facility issues have been largely neglected in research, policy, and practice.^3^

Robust epidemiological data for paediatric post-discharge mortality in the context of sepsis and severe infection have been limited. A growing evidence base clearly points to a significant burden of post-discharge mortality, which usually accounts for as many deaths as the acute hospital phase of illness.^4, 5^ While comorbid conditions such as malnutrition and anaemia have been clearly linked to risk, other factors such as illness severity (both at admission and discharge), prior hospitalizations, and underlying social vulnerability, have also been independently associated with poor post-discharge outcomes.^6^ However, we lack simple data-driven methods to identify those at highest risk of mortality.

Current epidemiological evidence has clearly demonstrated critical gaps in care following discharge.^7^ Most post-discharge deaths occur at home, rather than during a subsequent readmission, indicating poor health utilization among the most vulnerable. Effective health care utilization is often hampered by poverty, social dynamics within the community and the family, as well as poorly linked and unresponsive health facilities.^8–10^ The provision of quality care during and after discharge has also been shown to be a significant challenge in many facilities, at least in part due to severely strained human and material resources.

Thus, effective solutions to improving the transition of care from hospital to home within poorly resourced health systems must be child centred and focused on the identification of the most vulnerable children.^11^ In this study, we aim to update the development and validation of previously derived clinical prediction models that identify children, admitted with suspected sepsis, who are at risk of post-discharge mortality.^12^

## Methods

### Study design and approvals

Four independently funded, prospective observational cohort studies were conducted with a primary objective of generating data for model building: two were among children less than six months of age and two were among children 6-60 months of age.

These studies were approved by the institutional review boards of the Mbarara University of Science and Technology in Mbarara, Uganda (No. 15/10-16) and the University of British Columbia in Vancouver, Canada (H16-02679). This study was also approved by the Uganda National Council for Science and Technology (HS 2207). This manuscript adheres to the Transparent Reporting of a multivariable prediction model for Individual Prognosis Or Diagnosis (TRIPOD) statement.^13^

### Study setting and population

Subjects were enrolled from six hospitals in Uganda: the Mbarara Regional Referral Hospital (Southwestern Uganda), the Holy Innocents Children’s Hospital (Southwestern Uganda), the Uganda Ibanda Martyrs Hospital (Western Uganda), the Masaka Regional Referral Hospital (Central Uganda), the Villa Maria Hospital (Central Uganda) and the Jinja Regional Referral Hospital (Eastern Uganda) (see Supplementary Material S1 and Supplementary Table S1.1). These facilities serve the catchments of 30 districts with a total population of approximately 8·2 million individuals, including approximately 1·4 million children under five years of age.^14^ With a mix of urban and rural participants, this reflects a representative sampling of the Ugandan paediatric population.

Within each age cohort, all had identical eligibility criteria. All children who were admitted with suspected sepsis were eligible for enrolment. Suspected sepsis was defined as children who were admitted with a proven or suspected infection (as determined by the treating medical team). We have previously demonstrated that 90% of children enrolled using these criteria meet the international paediatric sepsis consensus conference (IPSCC) definition.^15^ The IPSCC defines sepsis as the presence of the systemic inflammatory response syndrome alongside a suspected or proven infection.

Enrolment in the first cohort occurred between 13 March 2012 and 13 January 2014 and was used in an earlier report of a predictive model for post-discharge mortality for children 6-60 months of age.^12^ The second and third cohorts, enrolled for this present analysis (primary study enrolment), were defined by their age ranges (0-6 months and 6-60 months) and have also been previously reported:^5^ 0-6-month-olds were enrolled between 11 January 2018 and 30 March 2020; 6-60-month-olds were enrolled between 13 July 2017 and 02 July 2019. The fourth cohort was enrolled only among children 0-6 months of age, for the purpose of understanding how the early period of the COVID-19 epidemic impacted post-discharge outcomes; this cohort enrolled children from 31 March 2020 until 05 August 2021. Protocols and procedures were largely overlapping, and the same group of research staff were involved in data collection during all four periods of enrolment.^16^

### Data collection procedures

All data collection tools are available through the Smart Discharges study Dataverse.^16^ Data collection procedures have been previously described (see also Supplementary Material S1).^5, 12^ Briefly, trained study nurses collected clinical, social, and demographic data from consented participants at the point of hospital admission. Though largely overlapping between the two age groups, some variables were specific to children less than 6 months. These variables were the candidate predictors for our models and were selected based on clinical and contextual knowledge of possible factors relating to post-discharge mortality. We used a modified Delphi process to identify the most promising candidate variables to be collected and used for model development in each age group.^17, 18^

At discharge, study nurses recorded discharge status (died, discharged, discharged against medical advice, referred) and discharge diagnosis. A field officer contacted enrolled children by phone at two and four months after discharge, and with an in-person visit at six months to determine mortality status and, if applicable, date of death. All data were collected using encrypted study tablets and then uploaded to a Research Electronic Data Capture (REDCap) database hosted at the BC Children’s Hospital Research Institute (Vancouver, Canada).^19, 20^

### Model development

#### Outcome definition and ascertainment

The primary outcome of the prediction model was post-discharge mortality within six months of discharge.

#### Variable selection

Recognising the challenges of implementing large prediction models in resource constrained settings, we determined *a priori* to develop three models for each age group (0-6-month-olds and 6-60-month-olds) and restricted each model to eight variables drawing from a different pool of available predictors: one model focused solely on commonly-available clinical variables; one model focused solely on commonly-available clinical and social variables; and one model that used any of the candidate predictor variables (**Figure 1**). A feature of our modelling approach (elastic net regression) is that the final model size could not be pre-specified, often resulting in large models. Therefore, we conducted two rounds of variable selection to achieve the desired model size.

**Figure 1.**
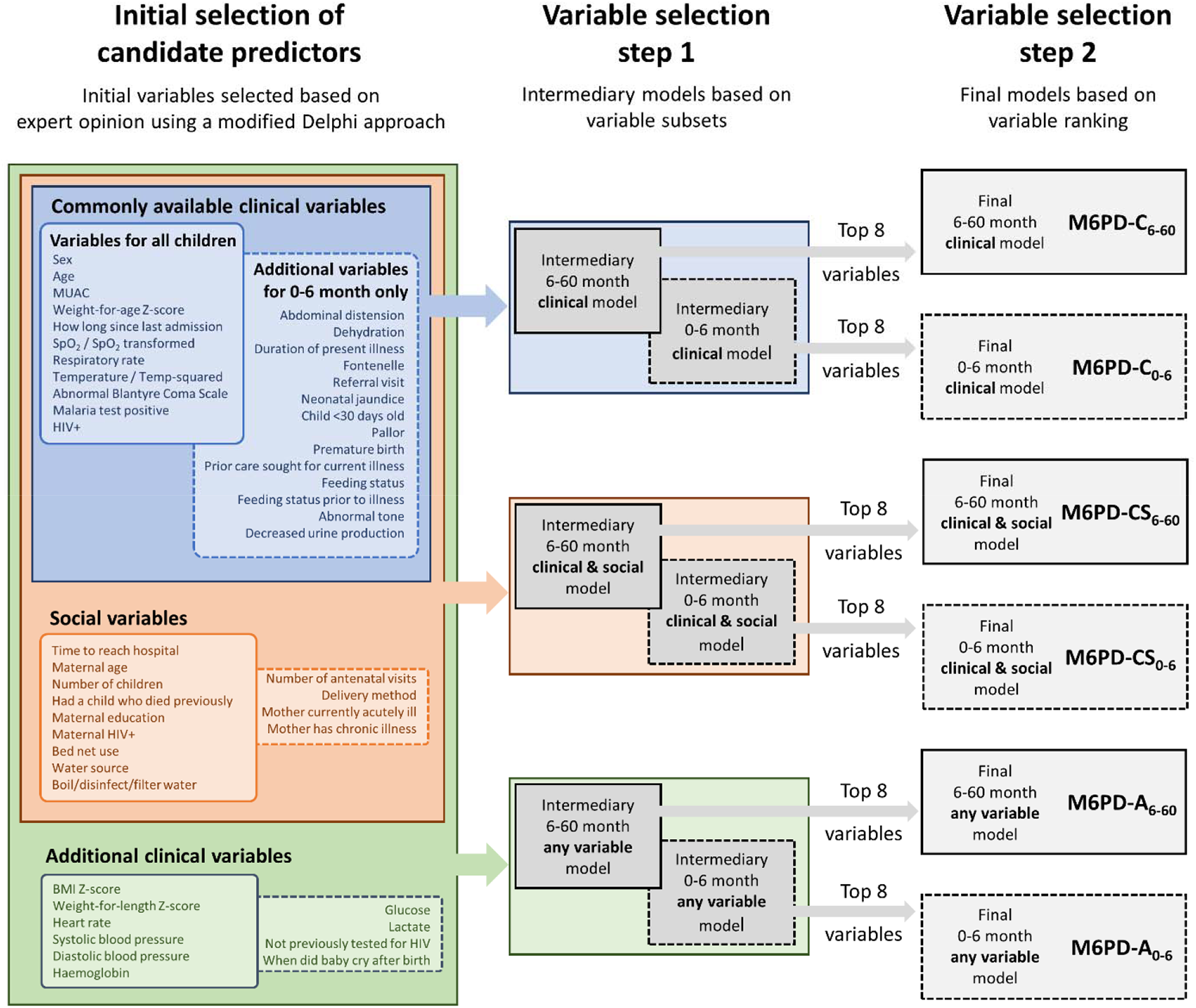
Variable selection for model development

To prioritize maximal parsimony, the first round of variable selection reduced the list of all possible candidate predictors to two smaller subsets: one subset including only the most relevant clinical variables; and a second subset including only the most relevant clinical and social variables. The variables included in these subsets were determined *a priori* by the investigators, based primarily on clinical significance and ease of measurement in low-resource settings. These subsets were used to derive the ***intermediary models*** that were either clinically-focused or clinically- and socially-focused; the full list of candidate predictor variables for each age group was also used to derive ***intermediary models*** that used any of the available variables. The full lists of candidate predictors used during the first round of variable selection in these ***intermediary models*** is given in Supplementary Tables S1.2 and S1.3.

The second round of variable selection involved calculating and ranking the importance of variables from each of the intermediary models: variable importance was calculated as the weighted sums of the absolute regression coefficients.^21^ The top eight unique variables (e.g., temperature and its quadratic term were considered as a single unique variable) were selected based on the average rank of 10-fold cross-validation of the intermediary models. If an interaction term was ranked as one of the top eight variables, then both terms in the interaction were included. The second round of variable selection produced a family of ***final models*** to predict mortality within six months post-discharge (M6PD) that used only the eight top-ranked variables in each age group: that is, models using only ***clinical*** variables, denoted by M6PD-C_0-6_ for the 0-6-month age group and M6PD-C_6-60_ for the 6-60-month age group; models using both ***clinical and social*** variables, denoted by M6PD-CS_0-6_ and M6PD-CS_6-60_; and models using ***any*** of the available predictor variables, denoted by M6PD-A_0-6_ and M6PD-A_6-60_ (Figure 1).

#### Adjustment and transformation of candidate predictors

All continuous variables were centred and scaled, and categorical variables converted to indicators (i.e. dummy variables). In addition to the raw oxygen saturation (SpO_2_) measurement, we used the transformation for SpO_2_ proposed by Zhou *et al.* to improve model prediction and calibration.^22^ Z-scored variables, including body mass index (BMI) z-scores, weight-for-age z-scores, and weight-for-length z-scores were calculated according to the World Health Organization (WHO) Child Growth Standards.^23^ Dehydration in the 0-6-month age group was determined using the WHO assessment criteria for dehydration.^24^ We included a quadratic term for temperature since both high and low temperatures may increase risk. Other nonlinearities were not considered due to risk of overfitting with the limited number of available events per variable.

### Statistical analysis

#### Sample size

The sample size for the primary study enrolment was determined to accomplish three primary aims. First, to explore the epidemiology of post-discharge mortality, which has been previously reported.^5^ Second, to develop prediction models. Third to act as a control period for a later interventional phase. For the present analysis, we determined the sample size required to develop a prediction model based on criteria proposed by Riley *et al*., 2020.^25^ For binary outcomes, three criteria are recommended based on: 1) reducing overfitting (caused by small sample sizes or too many candidate predictors relative to the sample size or number of events), defined by an expected shrinkage of predictor effects by ≤10%; 2) a small absolute difference of 0.05 in the apparent and adjusted Nagelkerke’s R^2^ value of the model, whereby the apparent R^2^ reflects the model performance in the same way that was used to develop the model and the adjusted R^2^ is an approximately unbiased estimate of the model fit;^26^ and 3) estimating the outcome proportion to within ±5% precision. The estimated sample size required to satisfy the three criteria was 2,117 and 1,551 for the 0-6-month and 6-60-month cohorts, respectively.

We also created *post hoc* learning curves of the sample size used to develop the model versus the area under the receiver operating characteristic curve (AUROC) when tested against a 20% hold-out set using the full set of variable predictors (Supplementary Figure S2.1).^27^ This involved building models using an increasing subset of the population (up to 80% of the total sample) and evaluating their performance against the hold-out set. For the 6-60-month learning curve, we started with the initial 1,242 children from 2012-2014,^12^ followed by the first 258 children recruited starting from July 2017 (n=1,500) and thereafter added groups of 500 consecutively recruited children until 80% (n=3,864) of the available population was reached. For the 0-6-month learning curve, we started with the first 500 children recruited starting from January 2018 and continued to add groups of 500 consecutively recruited children until 80% (n=2,679) of the available population was reached.

The performance of the 6-60-month derivation model increased with increasing sample size up to approximately 2,500 children after which the AUROC stabilized. The AUROC for the 0-6-month derivation model stabilized at approximately 1,000 children. This suggests that increasing our sample sizes beyond what we have currently collected would not result in further improvement of model performance.

#### Statistical methods and tools

All candidate predictors were summarised with means and standard deviations for continuous variables and counts and percentages for categorical variables. Kaplan-Meier survival curves were used to report the time to post-discharge death. As the amount of missing data overall and on any individual predictor was low, we used single-imputation with K-nearest neighbours imputation to replace missing values.^28^ Multiple imputation was considered, but not used, as this leads to additional complexity in the model building and validation steps.^29^

Elastic net regression was used to estimate coefficients for the prediction model.^30^ In sample sizes similar to ours, elastic net has been shown to perform similarly to more data-driven machine learning algorithms.^31, 32^ Optimal penalty terms were selected via 10-fold nested cross validation across a pre-specified grid of possible values. We developed full variable, intermediary, and final models utilizing the subset of variables as described in the previous section on model development.

Internal validation of the models was conducted using 10-fold cross validation of the model building process outlined above.^33^ This process consists of splitting the data into 10 ‘folds’, each one consisting of 90% of the original sample. The model is then built on each fold and tested on the held-out 10%. For each fold, we estimated the AUROC, the specificity, positive predictive value, and negative predictive value, using the probability threshold that gives 80% sensitivity, area under the precision recall curve, and Brier score. Internal model performance was assessed based on the cross-validated mean of the selected performance metrics. These performance metrics were also calculated on the entire dataset without cross-validation. We included additional plots for the gain curve, calibration, and the distribution of predicted probabilities stratified by mortality.

All analyses were conducted using R statistical software version 4.2.2 (R Foundation for Statistical Computing, Vienna, Austria) with the *caret* package (version 6.0-93) for model building and validation.^21, 34^

## Results

During the enrolment periods, a total of 22,166 consecutively admitted children were screened and 8,810 were enrolled (**Figure 2**). Among 0-6-month-olds (n=3,665), a total of 3,424 (93·4%) infants survived to discharge. Complete 6-month outcome data were available for 3,349 (97·8%) of these children, which formed the full dataset for model derivation and validation in this age group. Among 6-60-month-olds (n=5,145), a total of 4,916 (95·5%) children survived to discharge. Complete 6-month outcome data were available for 4,830 (98·2%) of these children, which formed the full dataset for model derivation and validation in this age group.

**Figure 2.**
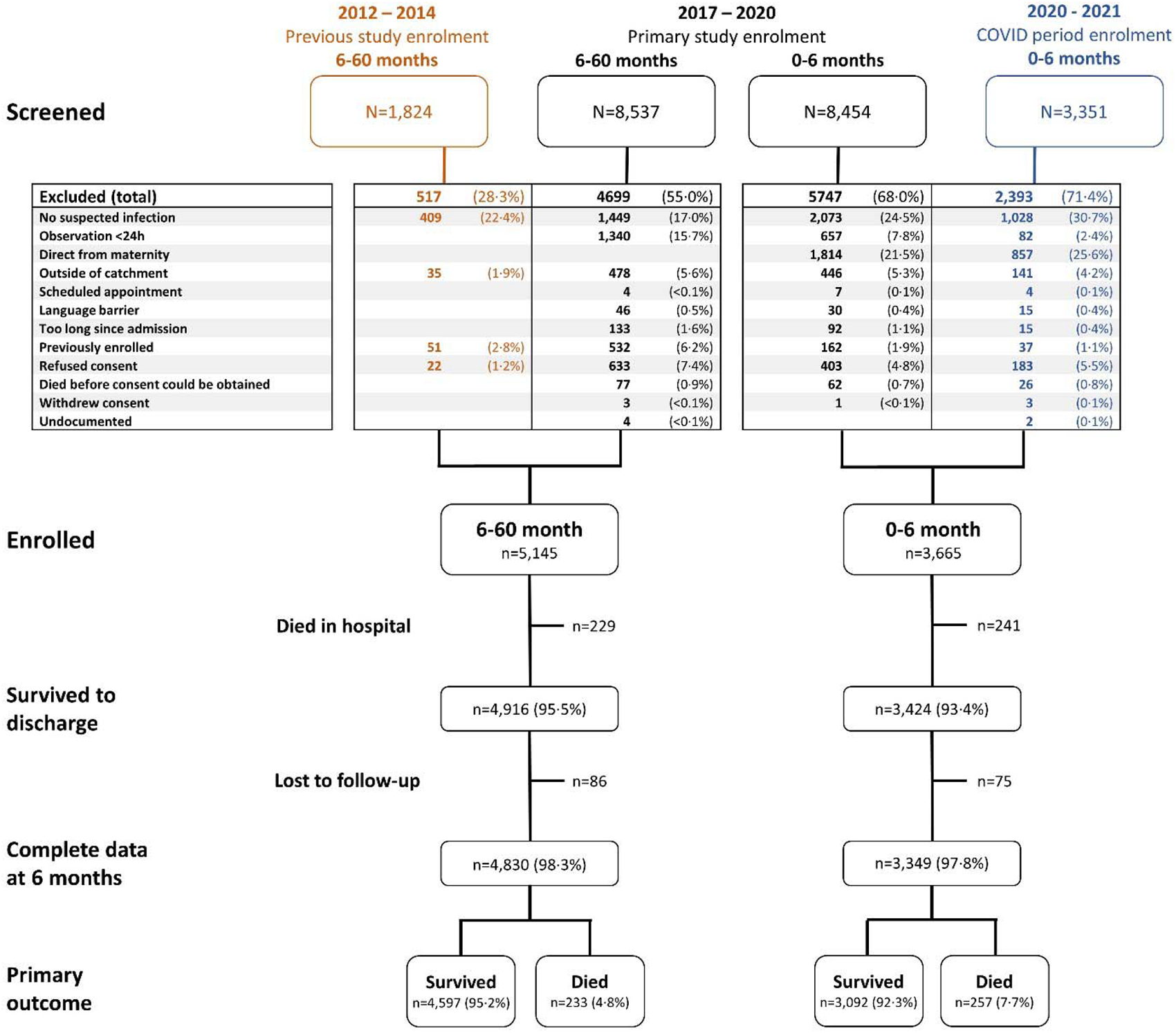
Study enrolment flow diagram

Mortality within 6 months of discharge occurred in 257 (7·7%) of those in the 0-6-month age group, with median (interquartile range [IQR]) time to death of 31 (9 – 80) days, and in 233 (4·8%) of the 6-60-month age group, with time to death of 36 (11 – 105) days (**Figure 3**). Missing data were minimal (**Table 1**, **Table 2**).

**Figure 3.**
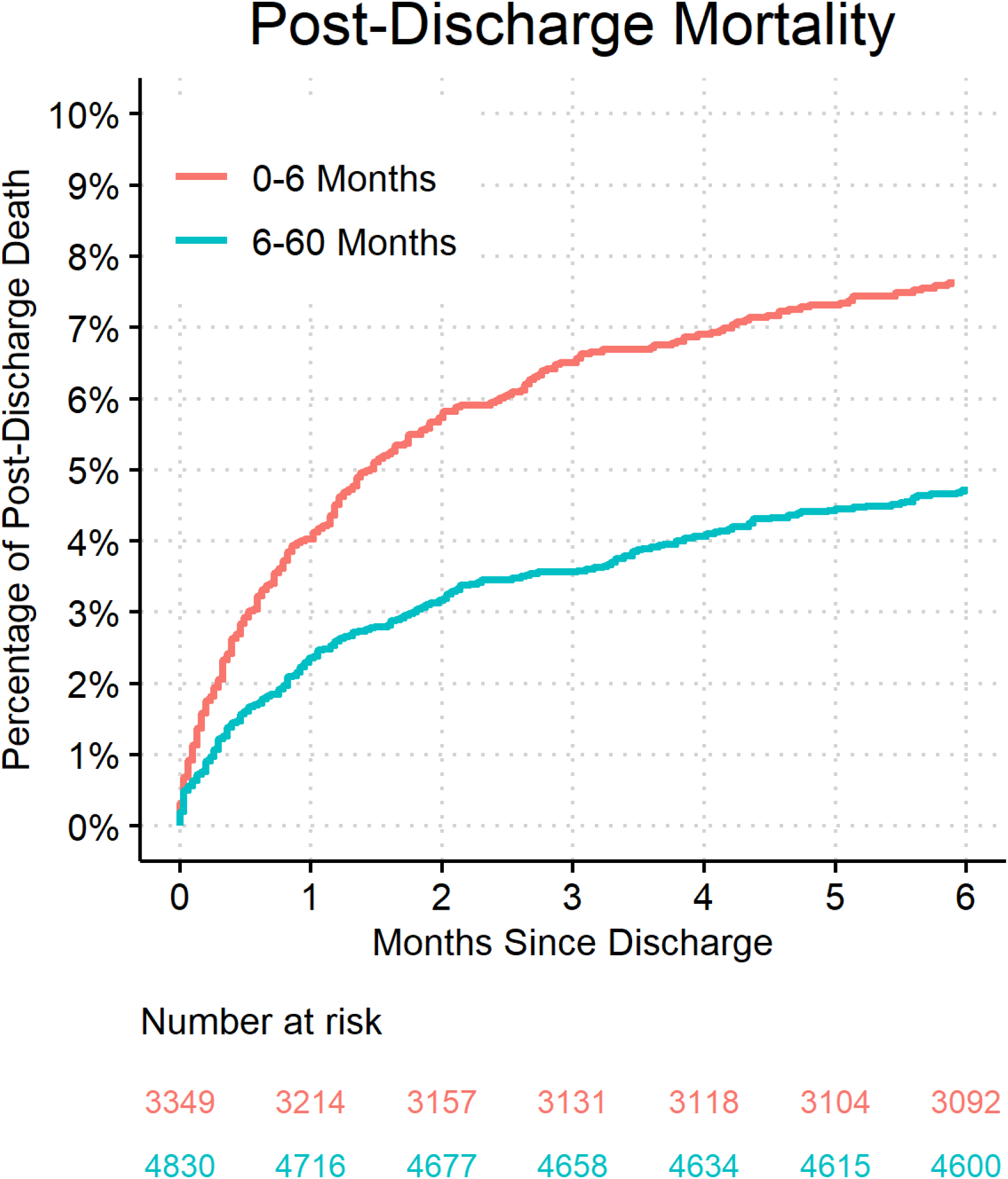
**Post-discharge mortality by age cohort.** Both the derivation and validation cohorts were combined for each of the age cohorts.

**Table 1.**
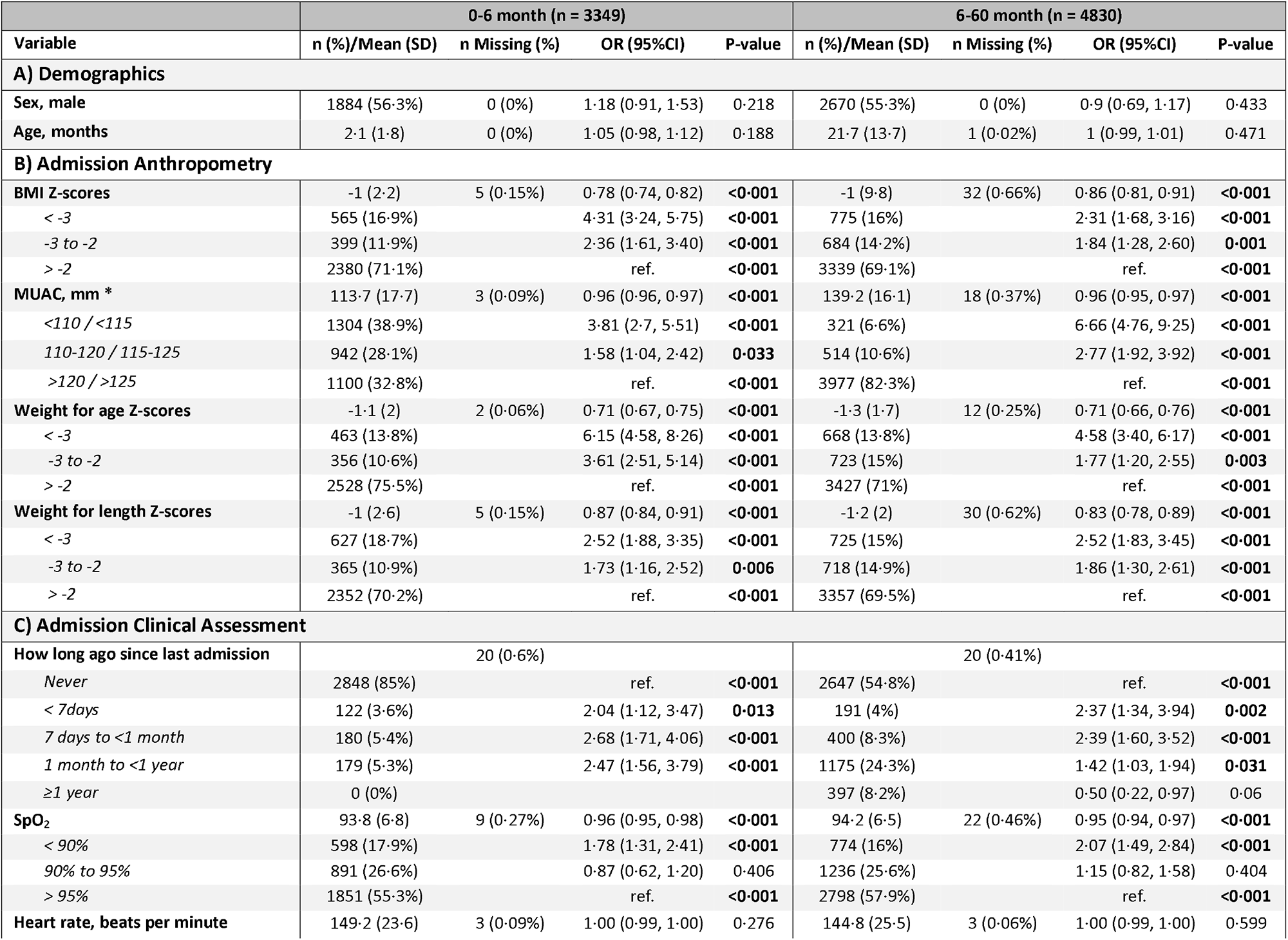
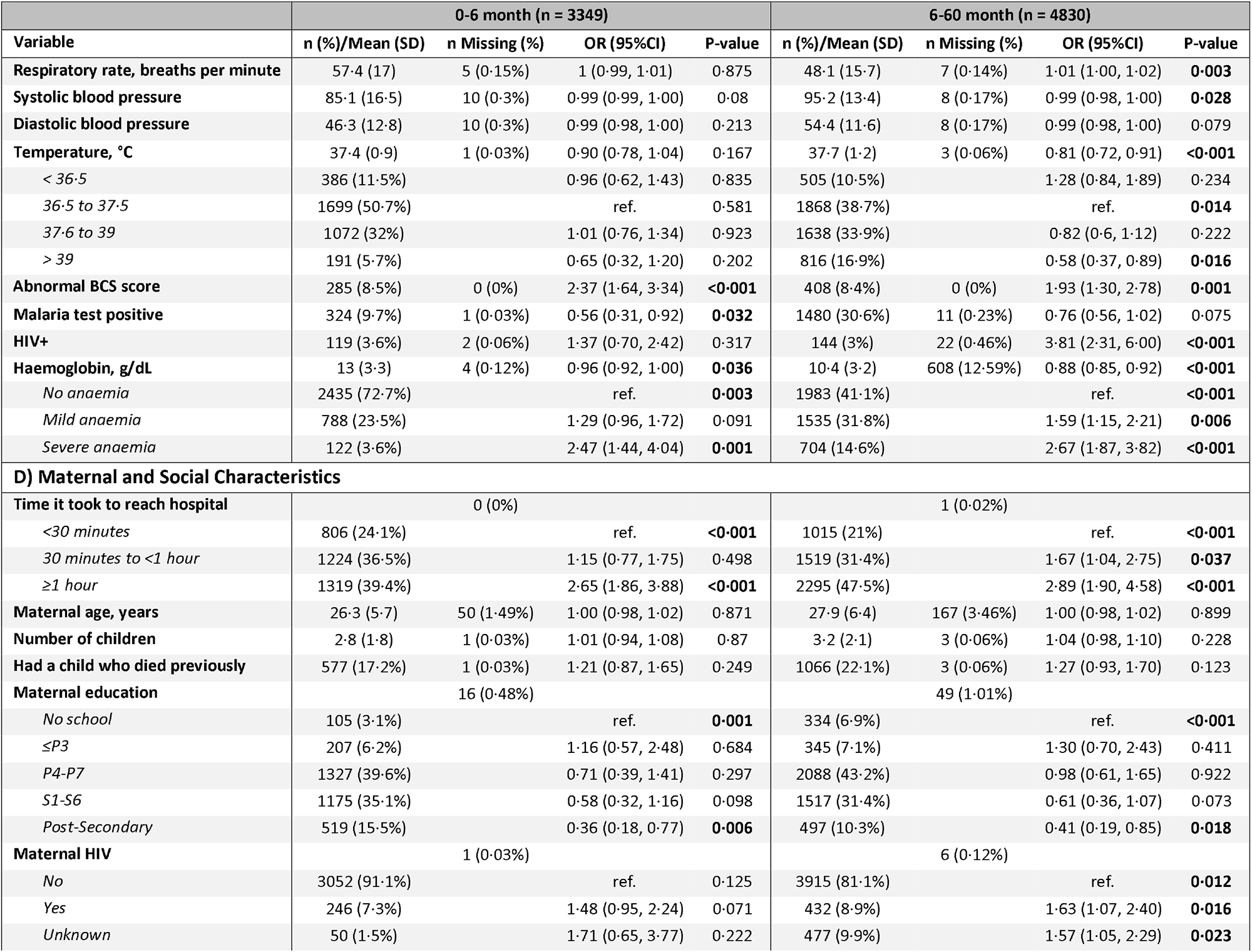
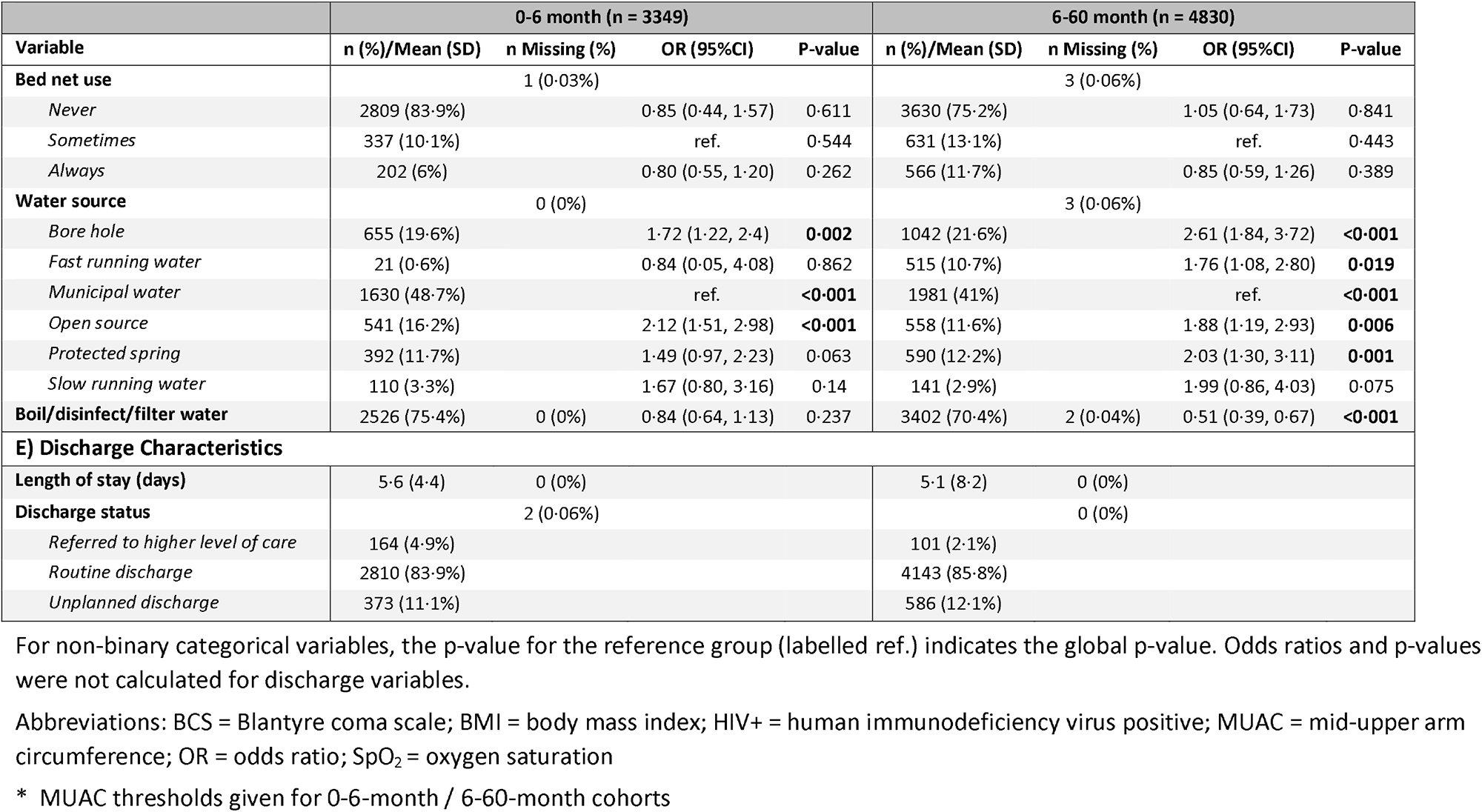
Demographics and univariable odds ratios for the risk of post-discharge infant mortality.

**Table 2.** Univariable odds ratios for post-discharge mortality for variables only collected among those <6m of age.

| Variable | n (%) / Mean (SD) | n Missing | OR (95%CI) | P-value |
| --- | --- | --- | --- | --- |
| **Abdominal distension** | 217 (6.5%) | 2 (0.06%) | 1.79 (1.15, 2.70) | **0.007** |
| **Antenatal visits** | 4.9 (1) | 48 (1.43%) | 0.89 (0.78, 1.01) | 0.066 |
| **Dehydration, WHO categories** |  | 11 (0.33%) |  |  |
| *No dehydration* | 2844 (84.9%) |  | ref. | **<0.001** |
| *Some dehydration* | 399 (11.9%) |  | 1.64 (1.15, 2.30) | **0.005** |
| *Severe dehydration* | 95 (2.8%) |  | 3.40 (1.96, 5.62) | **<0.001** |
| **Delivery method, caesarean** | 497 (14.8%) | 4 (0.12%) | 0.74 (0.49, 1.08) | 0.136 |
| **Duration of present illness** |  | 4 (0.12%) |  |  |
| <48 hours | 957 (28.6%) |  | ref. | **<0.001** |
| 48 hours to 7 days | 60 (1.8%) |  | 1.46 (1.05, 2.06) | **0.026** |
| 8 days to 1 month | 1985 (59.3%) |  | 3.16 (2.09, 4.80) | **<0.001** |
| >1 month | 343 (10.2%) |  | 5.13 (2.52, 9.87) | **<0.001** |
| **Fontanelle** | 132 (3.9%) | 6 (0.18%) | 2.4 (1.44, 3.81) | **<0.001** |
| **Glucose, mmol/L** | 5.7 (2.5) | 2 (0.06%) | 1.03 (0.98, 1.08) | 0.188 |
| **Not previously tested for HIV** | 2968 (88.6%) | 0 (0%) | 0.93 (0.64, 1.40) | 0.719 |
| **Referral visit** | 1056 (31.5%) | 1 (0.03%) | 1.70 (1.31, 2.20) | **<0.001** |
| **Neonatal jaundice** | 261 (7.8%) | 34 (1.02%) | 1.31 (0.83, 1.99) | 0.219 |
| **Lactate level, mmol/L** | 2.5 (1.6) | 9 (0.27%) | 1.10 (1.03, 1.18) | **0.003** |
| **Mother currently acutely ill** | 132 (3.9%) | 13 (0.39%) | 0.56 (0.22, 1.18) | 0.171 |
| **Mother has chronic illness** | 251 (7.5%) | 19 (0.57%) | 1.29 (0.81, 1.97) | 0.256 |
| **Child less than 30 days old** | 1353 (40.4%) | 2 (0.06%) | 0.68 (0.52, 0.89) | **0.006** |
| **Pallor** | 307 (9.2%) | 2 (0.06%) | 2.15 (1.50, 3.03) | **<0.001** |
| **Premature birth** | 210 (6.3%) | 6 (0.18%) | 2.05 (1.33, 3.06) | **0.001** |
| **Prior care sought for current illness** | 1995 (59.6%) | 0 (0%) | 1.82 (1.38, 2.42) | **<0.001** |
| **Sucking well when breastfeeding, or feeding well if not breastfed** | 1956 (58.4%) | 8 (0.24%) | 0.47 (0.36, 0.61) | **<0.001** |
| **Sucking well when breastfeeding, or feeding well if not breastfed, prior to illness** | 2589 (77.3%) | 392 (11.7%) | 0.59 (0.42, 0.85) | **0.004** |
| **When did the baby cry after birth** |  | 97 (2.9%) |  |  |
| *Immediately* | 2805 (83.8%) |  | ref. | **0.041** |
| <5 minutes | 138 (4.1%) |  | 1.83 (1.04, 3.02) | **0.025** |
| 5 to 10 minutes | 141 (4.2%) |  | 1.32 (0.70, 2.30) | 0.351 |
| 11 to 30 minutes | 68 (2%) |  | 1.26 (0.48, 2.72) | 0.594 |
| >30 minutes | 100 (3%) |  | 2.12 (1.14, 3.68) | **0.011** |
| **Abnormal tone** | 285 (8.5%) | 2 (0.06%) | 3.11 (2.21, 4.31) | **<0.001** |
| **Decreased urine production** | 677 (20.2%) | 99 (2.96%) | 1.91 (1.43, 2.52) | **<0.001** |

For non-binary categorical variables, the p-value for the reference group (labelled ref.) indicates the global p-value.
Abbreviations: CI = confidence interval; HIV = human immunodeficiency virus; OR = odds ratio; WHO = World Health Organization

Clinical and demographic details of these cohorts have been previously described (see also **Table 1**, **Table 2**).^5, 12^ The mean ±standard deviation [SD] age was 2·1 ±1·8 months in the 0-6-month age group and 21·7 ±13·7 months in the 6-60-month age group; 1,884 (56·3%) 0-6-month-olds were male and 2,670 (55·3%) 6-60-month-olds were male. Poor growth/malnutrition was common in both age groups, with 463 (13·8%) 0-6-month-olds and 668 (13·8%) 6-60-month-olds classified as severely underweight (weight for age z-score <-3) and similar weight-for-age z-score distribution in the two groups. Discharge diagnoses made by the clinical team were recorded and could be overlapping in the case of multiple diagnoses (Supplementary Table S1.4). Most variables considered in the modelling were associated with post-discharge mortality (**Table 1**, **Table 2**)

The intermediary variable models were large (see Supplementary Material S3 – S5). The models derived using all potential candidate predictors (intermediary any variable model) included 41 unique variables in the 0-6-month model and 19 unique variables in the 6-60-month model; coefficients, performance metrics, and variable importance are reported in Supplementary Material S5. When applied to the entire dataset for each age group, the AUROC was 0·81 (95%CI 0·79 to 0·84) for 0-6-month model and 0·79 (95%CI 0·77 to 0·82) for the 6-60-month model, with average AUROCs of 0·77 (range 0·69 to 0·87) and 0·76 (range 0·71 to 0·81) across the 10 cross-validations, respectively. Calibration was good in both age groups at low predicted probabilities, with a Brier score of 0·07 (range 0·06 to 0·07) for the 0-6-month model and 0·04 (range 0·04 to 0·05) for the 6-60-month models. Calibration decreased at higher predicted probabilities, although there were almost no individuals with probabilities exceeding 40%. In both age groups, mid-upper arm circumference (MUAC) was identified as the variable with the highest importance.

A summary of all final models is reported in **Table 3** and **Table 4**, and in detail in the Supplementary Material S6 – S8, which includes all model terms, their coefficients and variable importance plots outlining the relative importance of all coefficients in each of the models.

**Table 3.** Summary of performance and variables included in the set of final 0-6-month models with reduced number of variables using the probability threshold that gave a sensitivity of 0·8.

|  | **M6PD-C<sub>0-6</sub>**<br>**Model** | **M6PD-CS<sub>0-6</sub>**<br>**Model** | **M6PD-A<sub>0-6</sub>**<br>**Model** |
| --- | --- | --- | --- |
| **Average CV Performance** |  |  |  |
| Specificity | 0·60 | 0·61 | 0·61 |
| AUROC | 0·75 | 0·76 | 0·76 |
| PPV | 0·15 | 0·16 | 0·16 |
| NPV | 0·97 | 0·97 | 0·97 |
| PRAUC | 0·23 | 0·23 | 0·23 |
| Brier Score | 0·07 | 0·07 | 0·07 |
| **Full Dataset Performance** |  |  |  |
| Specificity | 0·58 | 0·62 | 0·62 |
| AUROC | 0·77 | 0·77 | 0·77 |
| PPV | 0·14 | 0·15 | 0·15 |
| NPV | 0·97 | 0·97 | 0·97 |
| PRAUC | 0·23 | 0·22 | 0·22 |
| Brier Score | 0·07 | 0·07 | 0·07 |
| **Variables** |  |  |  |
| Age, months | ✓ | ✓ | ✓ |
| Duration of present illness, categorical | ✓ | ✓ | ✓ |
| MUAC, mm | ✓ | ✓ | ✓ |
| Neonatal jaundice, binary | ✓ | ✓ | ✓ |
| Sucking well when breastfeeding, binary | ✓ | ✓ | ✓ |
| SpO<sub>2</sub>, % | ✓ | ✓ | ✓ |
| Time to reach hospital, categorical |  | ✓ | ✓ |
| Weight for age z-score | ✓ | ✓ | ✓ |
| Fontanelle, binary | ✓ |  |  |

Note, the final reduced and final clinical and social model are identical since the same variables were selected.
Abbreviations: AUROC = area under the receiver operating curve; MUAC = mid-upper arm circumference; NPV = negative predictive value; PPV = positive predictive value; PRAUC = area under the precision-recall curve; SpO<sub>2</sub> = oxygen saturation

**Table 4.**
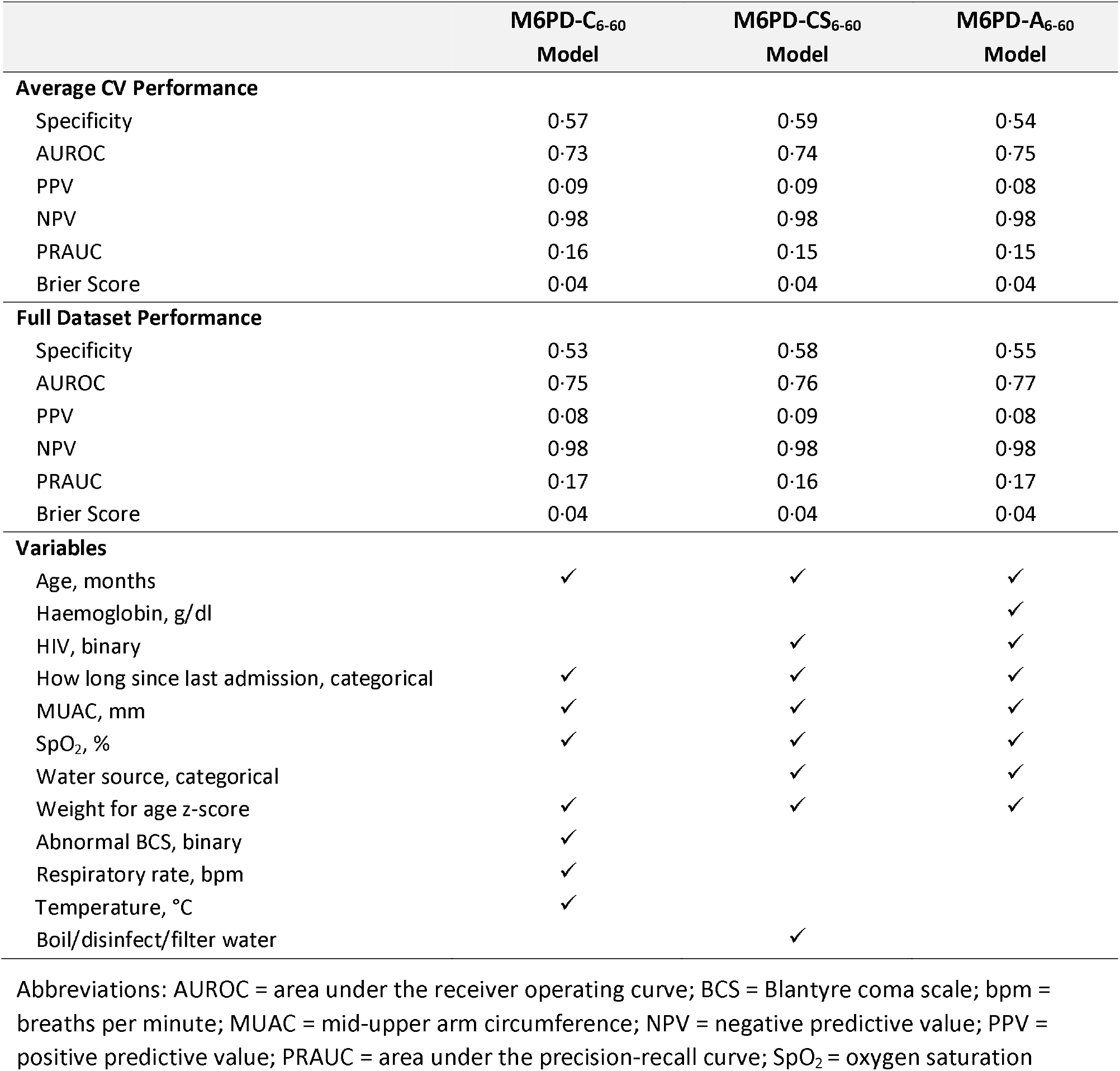
Summary of performance and variables included in the set of final 6-60-month models with reduced number of variables using the probability threshold that gave a sensitivity of 0·8.

The M6PD-C_0-6_ model, using only simple clinical variables, included weight-for-age z-score (mean rank 1·4, selection frequency 10), MUAC (mean rank 1·6, selection frequency 10), feeding status (mean rank 3·4, selection frequency 10), SpO_2_ (mean rank 5·8, selection frequency 9), duration of illness (mean rank 6·2, selection frequency 9), age × jaundice (mean rank 7·8, selection frequency 7), and bulging fontanelle (mean rank 8·3, selection frequency 8). This model had an AUROC of 0·77 (95%CI 0·74 to 0·80) when applied to the entire 0-6-month dataset (**Figure 4**), while the average AUROC across the internal 10 cross-validations was 0·75 (range 0·63 to 0·85). Setting the sensitivity to 80%, the corresponding probability threshold was 0·058; at this threshold, the positive and negative predictive values were 14% and 97% respectively. Calibration at low predicted probabilities was good, with a Brier score of 0·07. Calibration at probabilities beyond 30-40% was poor, but sample sizes were very small at this range.

**Figure 4.**
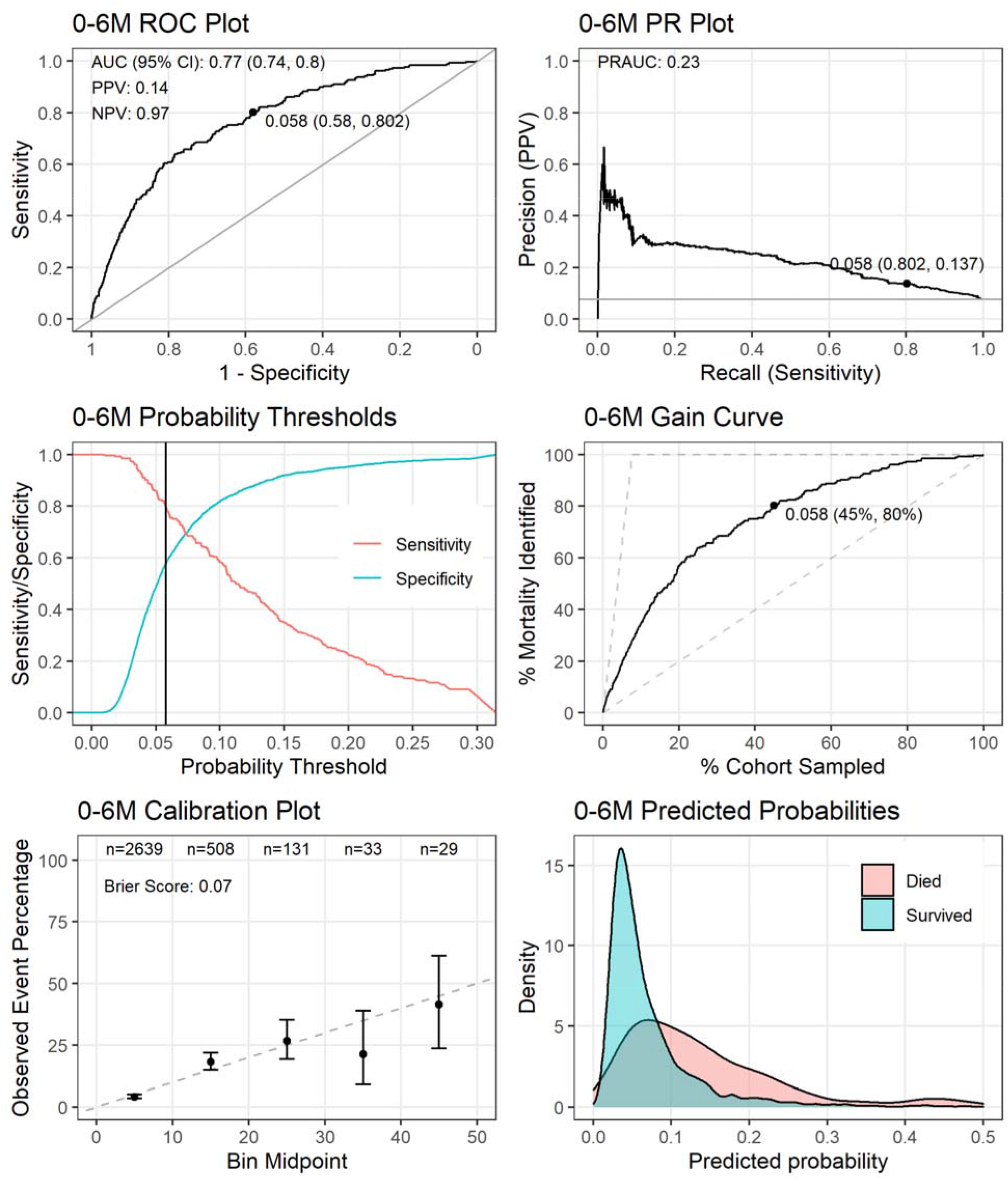
**Performance of the M6PD-C_0-6_ model** tested on the entire dataset after restricting the predictors to the eight unique variables from the intermediate clinical variable model with the highest average variable importance. The point on the receiver operating characteristic (ROC) plot, precision recall (PR) plot, and gain curve indicates the co-ordinates when using the probability threshold that gives a sensitivity of 80% (probability threshold = 0·058). The positive predictive value (PPV) and negative predictive value (NPV) are also reported in the ROC plot using this threshold.

The M6PD-CS_0-6_ model, using social and clinical variables, was nearly identical in performance to M6PD-C_0-6_; the variables were largely overlapping with only fontanelle status replaced by travel time required to reach hospital (**Table 3**; Supplementary Material S7). The M6PD-A_0-6_ model that used any available variable, was identical to M6PD-CS_0-6_ (**Table 3**; Supplementary Material S8).

The M6PD-C_6-60_ model, using only clinical predictors, included nine variables (the 8^th^ best performing variable included an interaction with a new variable; Supplementary Material S3). These variables included MUAC (mean rank 1, selection frequency 10), SpO_2_ (mean rank 2·7, selection frequency 10), weight-for-age z-score (mean rank 2·8, selection frequency 10), time since prior admission (mean rank 4·7, selection frequency 10), abnormal coma score (mean rank 5·8, selection frequency 9), temperature (mean rank 6·4, selection frequency 9), HIV status (mean rank 6·5, selection frequency 9) and age × respiratory rate (mean rank 9·1, selection frequency 2). This model had an AUROC of 0·74 (95% CI 0·72 to 0·79) when applied to the entire 6-60-month dataset (**Figure 5**), with an average AUROC of 0·73 (range 0·67 to 0·77) across the 10 cross-validations (**Table 4**; Supplementary Material S6). Setting sensitivity to 80%, the corresponding probability threshold was 0·036; at this threshold, the positive and negative predictive values were 0·08 and 0·98, respectively. Calibration across risk strata was good with a Brier score of 0·04.

**Figure 5.**
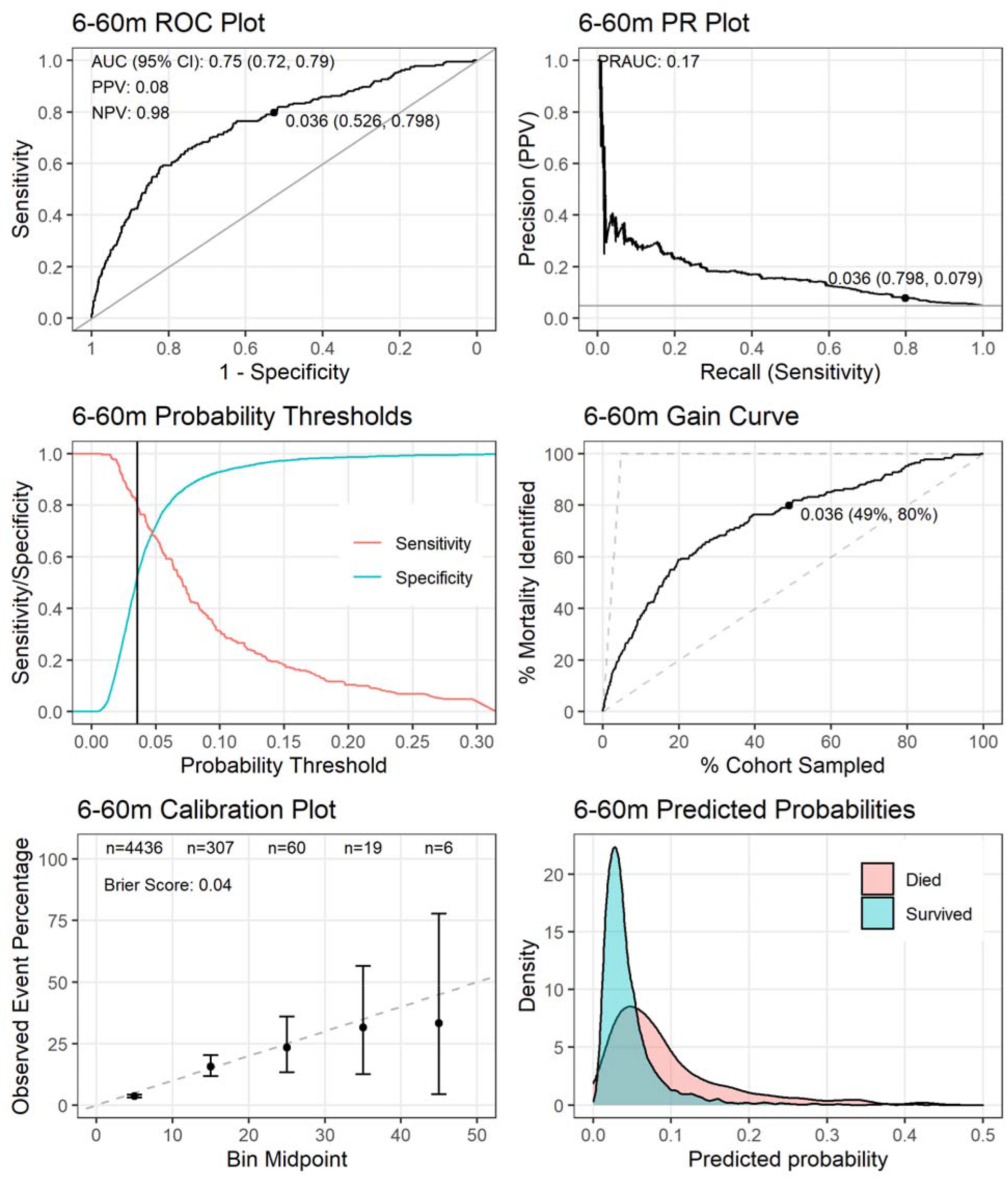
**Performance of the M6PD-C_6-60_ model** tested on the entire dataset after restricting the predictors to the nine unique variables from the intermediate clinical variable model with the highest average variable importance. The point on the receiver operating characteristic (ROC) plot, precision recall (PR) plot, and gain curve indicates the co-ordinates when using the probability threshold that gives a sensitivity of 80% (probability threshold = 0·036). The positive predictive value (PPV) and negative predictive value (NPV) are also reported in the ROC plot using this threshold.

The M6PD-CS_6-60_ model, which used clinical and social variables, performed nearly the same as M6PD-C_6-60_, with only home water source and water disinfection practices replacing coma score (**Table 4**; Supplementary Material S7). M6PD-A_6-60_ was similar to M6PD-CS_6-60_, except that water disinfection practices was replaced by haemoglobin; performance metrics were nearly identical (**Table 4**; Supplementary material S8).

## Discussion

Using four large, objective driven, prospective cohorts of under-5 children admitted with suspected sepsis, we derived and internally validated prediction models for post-discharge mortality using only admission data. The performance of these models to predict mortality out to 6 months post-discharge was good, suggesting their potential utility to link individual risk to the interventional intensity of a program to improve post-discharge outcomes among children.^35^ Data-driven, child-centred approaches to post-discharge care have been strongly advocated for recently.^4, 36, 37^ Utilizing data from multiple sites, captured over an 8-year timespan, we created robust, cross-validated models that should spur focus on external validation outside of a Ugandan context.

These results point not only to the high rates of post-discharge mortality, which have been previously demonstrated,^4, 5^ but also to the fact that those at highest risk can be reasonably identified using varied sets of simple and easy-to-collect variables. The development of multiple simplified models within each cohort may help alleviate the logistical barriers to implementation in different contexts. The required predictors are often routinely collected in similarly austere settings where such models could provide significant benefit. Notably, we observed similar between-model performance (within each cohort) and minimal reduction in performance in the intermediary models compared to the full models, which would themselves be logistically challenging to implement. Collinearity within the large sets of predictors may have accounted for this minimal loss in performance. Though our models do not accommodate missing data for predictor variables, the development of a family of models, varying in the number and identity of predictors, and producing similar performance, is possible, and could accommodate nearly every conceivable individual prediction scenario.

In the absence of an effective intervention, risk prediction has limited utility. An understanding of discharge as a dynamic process encompassing the time between admission and re-integration into community care is integral to our decision to focus on admission factors in model development. Early identification allows post-discharge risk to be incorporated into discharge planning from the earliest stages of admission. Prior work on understanding barriers to paediatric discharges have noted significant challenges related to preparing caregivers for discharge and the transition home, suggesting that early planning will be an essential component of effective peri-discharge care.^38^

Choosing risk probability thresholds to classify a binary outcome, such as post-discharge mortality, is dependent on many factors, including availability of human resources, baseline risk, risk tolerance, and impact on patients/caregivers. Though the risk classification thresholds chosen in this study may prove useful in some settings, the choice of both the threshold and the number of thresholds must be informed by the local context and constitutes a critically important consideration for the deployment of this, or any other, risk model.^39^

Although internal validation can justify the use of models within the region from which they are derived, external validation using different data sources (ideally several) for regions that differ from those where the models were derived is also important.^40^ We consider such validation exercises a high priority, using both existing and future data.^4^ To this end, we have several prospective studies currently underway, and plan to establish data sharing agreements with other collaborators to enable use of data collected by others. However, even under the most optimistic scenario not every conceivable implementation region for any given model will be subjected to external validation. A more pragmatic and relevant approach is therefore the development of a region-specific model updating process, integrated over the life-course of the model. Calibration drift due to secular trends, the measured impact of the model itself, and peculiarities of each individual site are key considerations in model deployment.^41^ Digitization of the healthcare system will aid in establishing these processes.^42^

As health systems in low-income country settings increasingly move towards the use of electronic health records, the incorporation of algorithms to augment care decisions has tremendous potential to both improve outcomes and to facilitate the adoption of these digital systems.^43, 44^ Use of routinely collected variables can allow models to run in the absence of any additional user input and thus automatically prompt follow-up guidance to both the medical team and the patient, encouraging adoption and linkage to interventional programs. Furthermore, use of such systems can also report baseline risk data and, when linked to follow-up programs, data on readmission and mortality to national-level health management information systems, such as DHIS2.^45^ These data can be used in model calibration and updating, ensuring site-specific validity.

This study is subject to several limitations. While our models performed reasonably well when assessed with cross validation, it remains to be seen how well they perform under external validation. Demonstrating good external performance will be helpful to encourage adoption, and will occur in due course. Second, these models were developed in the absence of a proven program to utilize a risk-based approach to care, limiting their current utility. While merely knowledge of individual risk can change behaviour and may influence the provision of discharge and post-discharge care, risk-informed approaches to follow-up care are also currently under investigation and will be reported once complete.^46^ Third, calibration was good at most observed risks but there were very few patients with predicted risks greater than 40-50% so calibration beyond these risk probabilities could not be measured adequately. Regardless, our models should perform well for implementation purposes using the optimal threshold cut-offs identified. Finally, the added value of these models may be questioned in the light of previously developed and published models.^46–48^ However, our models were based on purposively built cohorts, with *a priori* stakeholder engagement regarding relevant variables and their measurement timing. Furthermore, these models were uniquely developed within the clinical rubric of suspected sepsis, which is increasingly recognized as a global health priority.

Post-discharge mortality in the context of suspected sepsis occurs frequently, but those at highest risk can be identified using simple clinical criteria, measured at admission. Though future external validation is required, the use of these models within the context where they were collected can begin to transform discharge and post-discharge care.

## Data Availability

Study materials (including de-identified data, protocol, data collection tools and analysis code) are available upon reasonable request to the corresponding author or through the Pediatric Sepsis CoLab. The University of British Columbia Dataverse Collection: Pediatric Sepsis CoLab. Smart Discharges Dataverse. Borealis. 2022. https://borealisdata.ca/dataverse/smart_discharge

## Funding

This study was funded by Grand Challenges Canada (grant #TTS-1809-1939), the Thrasher Research Fund (grant #13878), the BC Children’s Hospital Foundation, and Mining4Life.

We would like to acknowledge all past and present members of the Smart Discharges Research program for their efforts in data collection, administration, logistics support, and all study activities, including but not limited to: Tumwebaze Godfrey, Agaba Collins, Tumukunde Goreth, Naturinda Mackline, Assimwe Abibu, Nakafero Joan, Kiiza Israel, Kitenda Julius, Kamba Ayub, Kuguminkiriza Brenda, Kabajasi Olive, Kembabazi Brenda, Happy Annet, Tusingwire Fredson, Nuwasasira Agaston, Ankatse Christine, Naturinda Rabecca, Nabawanuka Abbey Onyachi, Kamazima Justine, Kairangwa Racheal, Ounyesiga Thomas, Mwoya Yuma, Twebaze Florence, Bulage Mary, Tugumenawe Darius, Tuhame Dyonisius, Twesigye Leonidas, Kamusiime Olivia, Ainembabazi Harriet, Abaho Samuel, Nakabiri Zaituni, Naigaga Shaminah, Kisame Zorah, Babirye Clare, Kayegi Maliza, Opuko Wilson, Mwaka Savio, Baryahirwa Hassan, Mutungi Alexander, Charlene Kanyali, Catherine Kiggundu, Alexia Krepiakevich, Brooklyn Nemetchek, Jessica Trawin, Maryum Chaudhry, Peter Lewis, Rishika Bose, Sahar Zandi Nia, Tamara Dudley. Without their effort and support, this study would not have been possible.

## Author contributions

Contributed to funding applications: MOW, JNB, EK, AT, CB, JS, CO, JMA, NK, JS, CPL, PML, PPM, SN, NKM, JN, JK. Designed the study: MOW, EK, AT, CB, JS, CO, JMA, NK, JS, CPL, PML, PPM, NKM, JK, DD. Coordinated study implementation: MOW, EK, CK, MT, DM, JK. Supervised data collection: SB, AT, SOS, EB, ES, CB, DD, CK, MT, DM. Managed the data: MOW, DD, CK, MT, DM, CZ, MK. Analysed the data: MOW, VN, JNB, CZ, MK, CK. Interpreted the data: MOW, VN, JNB, EK, AT, JMA, NK, JS, CPL, PML, PPM, SN, NKM, NW, JN, JK. Drafted the manuscript: MOW, VN, NW. Critically reviewed the manuscript: MOW, VN, JNB, EK, SB, AT, SOS, EB, ES, CB, JN, CO, JMA, NK, JS, CPL, PML, DD, PPM, SN, CK, MT, DM, CZ, MK, NW, NKM, JK. All authors had full access to all the data in the study, which was verified by MOW, VN, JNB, CK, JK, DD, and CZ; all authors have reviewed and approved the final manuscript text, and accept responsibility to submit for publication.

