## Supplementary material for "Prediction models for post-discharge mortality among under-five children with suspected sepsis in Uganda: A multicohort analysis"

### S1: Details of study cohorts and variables used for the full and intermediary models

#### Recruitment details, timelines and sites

The overall cohort of children 6-60 months of age consisted of a previously collected and reported dataset of 1307 children, of whom 1242 were discharged alive,^1^ and data collected for the present study between July 2017 and July 2020;^2^ both studies used identical eligibility criteria. The main cohort of children 0-6 months of age were recruited between January 2018 and March 2020, along with an additional cohort enrolled between March 2020 and August 2021. All children were followed up for 6 months after hospital discharge.

Children who resided outside of the hospital catchment, or were admitted for a brief period of observation, were excluded. Children admitted immediately after birth (i.e., without having first been discharged) were also excluded from the 0-6-month cohort.

All study materials are available in the Smart Discharges study Dataverse, including: the study protocol and data collection tools;^3^ and the data, analysis code, and data dictionary.^4^

1	Wiens MO, Kumbakumba E, Larson CP, *et al.* Postdischarge mortality in children with acute infectious diseases: derivation of postdischarge mortality prediction models. *BMJ Open* 2015; **5**: e009449.

2	Wiens MO, Bone J, Kumbakumba E, *et al.* Mortality after hospital discharge among children younger than 5 years admitted with suspected sepsis in Uganda: a prospective, multisite, observational cohort study. *Lancet Child Adolsecent Heal* 2023; : In press (accepted 01Mar2023).

3	Wiens M, Kissoon N (Tex), Ansermino JM, *et al.* Smart Discharges to improve post-discharge health outcomes in children: A prospective before-after study with staggered implementation. Borealis, V1. 2023. <https://doi.org/10.5683/SP3/QRUMNQ> (accessed May 2, 2023).

4	Wiens MO, Bone JN, Kumbakumba E, *et al.* Post-discharge mortality among children under 5 years admitted with suspected sepsis in Uganda: a prospective multi-site study ~ Smart Discharges. Borealis, V2. 2022. <https://doi.org/10.5683/SP3/REPMSY> (accessed Dec 19, 2022).

#### **Table S1.1** Study enrolment sites

| **Enrolment period** | **2012 – 2013** | **2017-2019** | **2018-2020** | **2020 – 2021** |
| --- | --- | --- | --- | --- |
| **Cohort** | **6-60-month** | **6-60-month** | **0-6-month** | **0-6-month** |
| **Mbarara Regional Referral Hospital**  (Southwestern Uganda) | ✓ | ✓ | ✓ | ✓ |
| **Holy Innocents Children’s Hospital**  (Southwestern Uganda) | ✓ | ✓ | ✓ | ✓ |
| **Masaka Regional Referral Hospital**  (Central Uganda) |  | ✓ | ✓ | ✓ |
| **Jinja Regional Referral Hospital**  (Eastern Uganda) |  | ✓ | ✓ | ✓ |
| **Villa Maria Hospital**  (Central Uganda) |  |  | ✓ | ✓ |
| **Uganda Martyrs Hospital, Ibanda**  (Southwestern Uganda) |  |  | ✓ | ✓ |

#### **Table S1.2** Candidate variables used in the full prediction model, intermediary clinical variable model, and the intermediary clinical and social variable model for both **0-6-month** and **6-60-month** cohorts. See Table S1.3 for additional variables used only in the 0-6-month cohort.

| **Variable** | **Full Model** | **Intermediary Clinical Variable Model** | **Intermediary Clinical and Social Variable Model** |
| --- | --- | --- | --- |
| Sex, binary | ✓ | ✓ | ✓ |
| Age, months | ✓ | ✓ | ✓ |
| BMI Z-score | ✓ |  |  |
| MUAC, mm | ✓ | ✓ | ✓ |
| Weight for age Z-score | ✓ | ✓ | ✓ |
| Weight for length Z-score | ✓ |  |  |
| How long since last admission, categorical | ✓ | ✓ | ✓ |
| SpO_2,_ % | ✓ | ✓ | ✓ |
| SpO_2_ transformed | ✓ | ✓ | ✓ |
| Heart rate, beats per minute | ✓ |  |  |
| Respiratory rate, breaths per minute | ✓ | ✓ | ✓ |
| Systolic blood pressure, mmHg | ✓ |  |  |
| Diastolic blood pressure, mmHg | ✓ |  |  |
| Temperature, °C | ✓ | ✓ | ✓ |
| Temperature-squared | ✓ | ✓ | ✓ |
| Abnormal BCS, binary | ✓ | ✓ | ✓ |
| Malaria test positive, binary | ✓ | ✓ | ✓ |
| HIV+, binary | ✓ | ✓ | ✓ |
| Haemoglobin, g/dl | ✓ |  |  |
| Time to reach hospital, categorical | ✓ |  | ✓ |
| Maternal age, years | ✓ |  | ✓ |
| Number of children | ✓ |  | ✓ |
| Had a child who died previously, binary | ✓ |  | ✓ |
| Maternal education, categorical | ✓ |  | ✓ |
| Maternal HIV, categorical | ✓ |  | ✓ |
| Bed net use, categorical | ✓ |  | ✓ |
| Water source, categorical | ✓ |  | ✓ |
| Boil/disinfect/filter water, binary | ✓ |  | ✓ |

Abbreviations: BCS = Blantyre coma scale; HIV = human immunodeficiency virus; MUAC = mid-upper arm circumference; SpO_2_ = oxygen saturation

#### **Table S1.3** List of candidate variables used in the full prediction model, intermediary clinical variable model, and the intermediary clinical and social variable model for only the **0-6-month** cohort

| **Variable** | **Full Model** | **Intermediary Clinical Variable Model** | **Intermediary Clinical and Social Variable Model** |
| --- | --- | --- | --- |
| Abdominal distension, binary | ✓ | ✓ | ✓ |
| Number of antenatal visits | ✓ |  | ✓ |
| Dehydration, WHO categories | ✓ | ✓ | ✓ |
| Delivery method, binary | ✓ |  | ✓ |
| Duration of present illness, categorical | ✓ | ✓ | ✓ |
| Fontenelle, binary | ✓ | ✓ | ✓ |
| Glucose, mmol/L | ✓ |  |  |
| Not previously tested for HIV, binary | ✓ |  |  |
| Referral visit, binary | ✓ | ✓ | ✓ |
| Neonatal jaundice, binary | ✓ | ✓ | ✓ |
| Lactate, mmol/L | ✓ |  |  |
| Mother currently acutely ill, binary | ✓ |  | ✓ |
| Mother has chronic illness, binary | ✓ |  | ✓ |
| Child less than 30 days old, binary | ✓ | ✓ | ✓ |
| Pallor, binary | ✓ | ✓ | ✓ |
| Premature birth, binary | ✓ | ✓ | ✓ |
| Prior care sought for current illness, binary | ✓ | ✓ | ✓ |
| Sucking well when breastfeeding, or feeding well if not breastfed, binary | ✓ | ✓ | ✓ |
| Sucking well when breastfeeding, or feeding well if not breastfed, prior to illness, binary | ✓ | ✓ | ✓ |
| When did baby cry after birth, categorical | ✓ |  |  |
| Abnormal tone, binary | ✓ | ✓ | ✓ |
| Decreased urine production, binary | ✓ | ✓ | ✓ |

Abbreviations: HIV = human immunodeficiency virus; WHO = World Health Organization

#### **Table S1.4** Diagnoses at discharge

|  | **0-6-month cohort, n (%)**  **N=3,349** | **6-60-month cohort, n (%)**  **N=4,830** |
| --- | --- | --- |
| Bronchiolitis | 233 (7%) | 158 (3·3%) |
| Diarrhoea/gastroenteritis | 262 (7·8%) | 842 (17·4%) |
| Febrile convulsions | 6 (0·2%) | 23 (0·5%) |
| HIV or HIV related illness | 20 (0·6%) | 98 (2%) |
| Malaria | 287 (8·6%) | 1711 (35·4%) |
| Malnutrition | 131 (3·9%) | 393 (8·1%) |
| Measles | 56 (1·7%) | 376 (7·8%) |
| Meningitis/encephalitis | 155 (4·6%) | 118 (2·4%) |
| Pneumonia | 1130 (33·7%) | 1489 (30·8%) |
| Reactive airway disease/asthma | 6 (0·2%) | 33 (0·7%) |
| Respiratory tract infection (cold, flu, etc.) | 131 (3·9%) | 420 (8·7%) |
| Sepsis | 1479 (44·2%) | 818 (16·9%) |
| Skin or soft tissue infection | 166 (5%) | 68 (1·4%) |
| Tuberculosis | 17 (0·5%) | 86 (1·8%) |
| Other infectious disease | 61 (1·8%) | 119 (2·5%) |
| Other non-infectious disease | 319 (9·5%) | 202 (4·2%) |

### S2: Learning Curve of Training Sample Size vs Performance

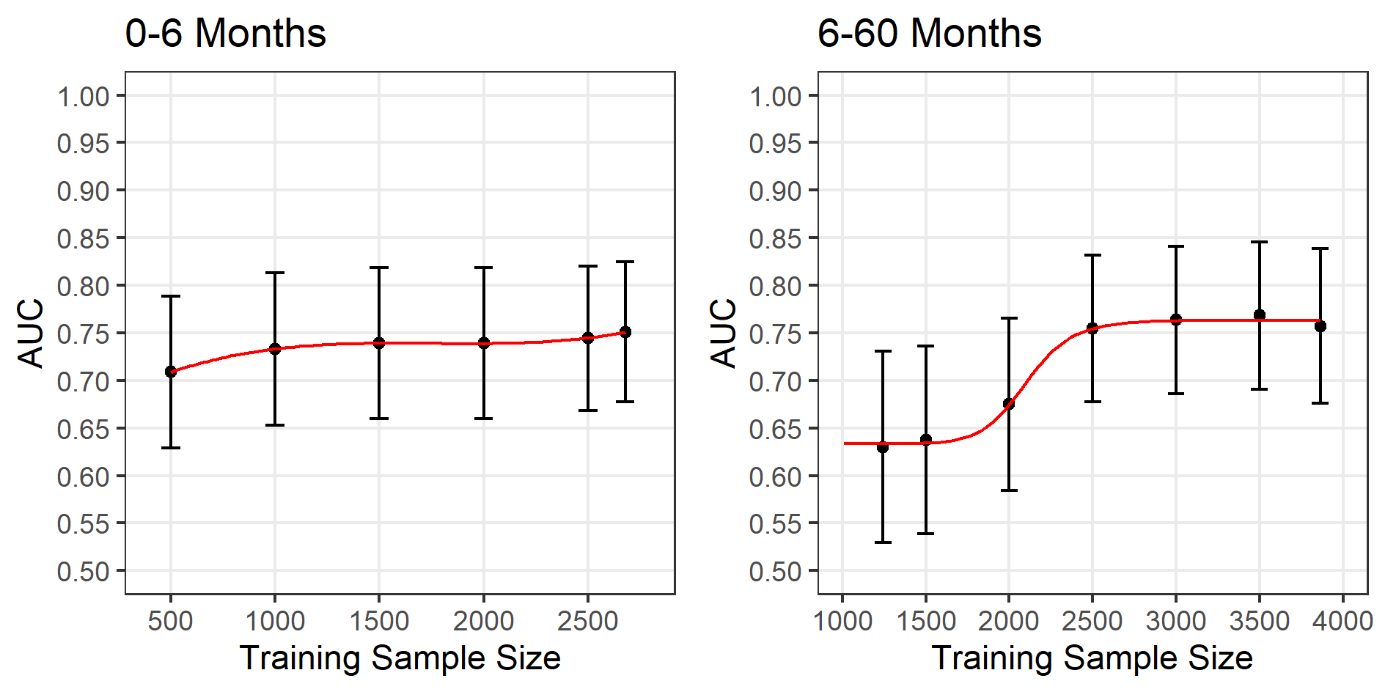

#### **Figure S2.1.** Learning curve showing the performance in terms of area under the curve (AUC) and 95% confidence intervals of the elastic net derivation model against the training sample size.

Performance was evaluated using the last 20% of children that were recruited for each age cohort. The training samples were increased based on chronological order of recruitment, i.e., for the **0-6-month** cohort, the first 500 patients recruited in the study were used to create the model and its performance tested on the 20% holdout dataset. This was repeated sequentially until the remaining 80% of the cohort was used. The training sample for the **6-60-month** cohort started with the initial 1307 patients recruited in 2012-2013.

### S3: Intermediary Clinical Variable Models – Variable Importance

#### **Table S3.1** Average rank of variable importance and the number of times selected in the top 8 variables across 10 folds of cross-validation from the **0-6-month** intermediary clinical variable model.

Only the top 20 variables and interactions are shown. Interactions between variables are indicated by the multiplication sign. The top eight unique variables (highlighted in bold) by average rank were used in the final model including their interactions with age.

| **Variable** | **Average Rank** | **Times Selected in Top 8** |
| --- | --- | --- |
| **Weight for age z-score** | **1·4** | **10** |
| **MUAC** | **1·6** | **10** |
| **Sucking well when breastfeeding, or feeding well if not breastfed** | **3·4** | **10** |
| **SpO_2_** | **5·8** | **9** |
| **Duration of present illness, 8 days – 1 month** | **6·2** | **9** |
| **Age × Jaundice** | **7·8** | **7** |
| **Fontanelle** | **8·3** | **8** |
| Neonate | 8·7 | 5 |
| Prior care sought for current illness | 11·3 | 2 |
| Age × Sucking well when breastfeeding, or feeding well if not breastfed, prior to illness | 11·6 | 5 |
| Malaria | 11·9 | 0 |
| Abnormal tone | 12·5 | 2 |
| Visit referred | 13·0 | 1 |
| Age × How long since last admission, <7 days | 13·6 | 0 |
| Pallor | 16·6 | 0 |
| Abdominal tension | 17·4 | 0 |
| Decreased urine production | 18·7 | 0 |
| How long since last admission, 1 month – 1 years | 19·5 | 0 |
| How long since last admission, 7 days – 1 month | 21·6 | 0 |
| Age × SpO_2_ | 24·7 | 0 |

Abbreviations: MUAC = mid-upper arm circumference; SpO_2_ = oxygen saturation

#### **Table S3.2** Average rank of variable importance and the number of times selected in the top 8 variables across 10 folds of cross-validation from the **6-60-month** intermediary clinical variable model.

Only the top 20 variables and interactions are shown. Interactions between variables are indicated by the multiplication sign. The top nine^1^ unique variables (highlighted in bold) were used in the final model including their interactions with age.

| **Variable** | **Average Rank** | **Times Selected in Top 8** |
| --- | --- | --- |
| **MUAC** | **1·0** | **10** |
| **SpO_2_** | **2·7** | **10** |
| **Weight for age z-score** | **2·8** | **10** |
| **How long since last admission, 7 days – 1 month** | **4·7** | **10** |
| **Abnormal BCS** | **5·1** | **9** |
| **Temperature, °C** | **6·4** | **9** |
| **HIV+** | **6·5** | **9** |
| **Temperature-squared, °C** | **8·0** | **6** |
| **Age × Respiratory rate** | **9·1** | **2** |
| Age × How long since last admission, 1 month – 1 year | 10·5 | 3 |
| How long since last admission, >1 year | 11·3 | 0 |
| How long since last admission, <7 days | 11·4 | 1 |
| Respiratory rate | 13·7 | 1 |
| Age × Abnormal BCS | 16·5 | 0 |
| Age × How long since last admission, <7 days | 17·4 | 0 |
| Age | 18·2 | 0 |
| How long since last admission, 1 month – 1 year | 18·2 | 0 |
| Age × How long since last admission, 7 days – 1 month | 19·4 | 0 |
| Age × HIV+ | 21·4 | 0 |
| Age × MUAC | 21·9 | 0 |

^1^ Nine variables were selected here as the eighth unique variable with the highest importance was within an interaction effect.

Abbreviations: BCS = Blantyre coma scale; HIV = human immunodeficiency virus; MUAC = mid-upper arm circumference; SpO_2_ = oxygen saturation

### S4: Intermediary Clinical and Social Variable Models – Variable Importance

#### **Table S4.1** Average rank of variable importance and the number of times selected in the top 8 variables across 10 folds of cross-validation from the **0-6-month** intermediary clinical and social variable model.

Only the top 20 variables and interactions are shown. Interactions between variables are indicated by the multiplication sign. The top eight unique variables (highlighted in bold) by average rank were used in the final model including their interactions with age.

| **Variable** | **Average Rank** | **Times Selected in Top 8** |
| --- | --- | --- |
| **Weight for age z-score** | **1·1** | **10** |
| **MUAC** | **1·9** | **10** |
| **Time it took to reach hospital, >1 hour** | **3·2** | **10** |
| **Sucking well when breastfeeding, or feeding well if not breastfed** | **4·5** | **10** |
| **SpO_2_** | **6·3** | **8** |
| **Duration of present illness, 8 days – 1 month** | **7·4** | **7** |
| **Age × Neonatal jaundice** | **9·4** | **4** |
| Abnormal tone | 9·7 | 4 |
| Fontanelle | 10·3 | 7 |
| Prior care sought for current illness | 15·0 | 0 |
| Age × How long since last admission, <7 days | 15·0 | 1 |
| Malaria | 15·3 | 1 |
| Neonate | 15·9 | 1 |
| Maternal education | 16·6 | 0 |
| Decreased urine production | 19·7 | 0 |
| Water source, open source | 19·9 | 0 |
| Pallor | 20·2 | 0 |
| Age × Weight for age z-score | 21·0 | 5 |
| How long since last admission, 1 month – 1 year | 21·1 | 0 |
| How long since last admission, 7 days – 1 month | 22·6 | 0 |

Abbreviations: MUAC = mid-upper arm circumference; SpO_2_ = oxygen saturation

#### **Table S4.2** Average rank of variable importance and the number of times selected in the top 8 variables across 10 folds of cross-validation from the **6-60-month** intermediary clinical and social variable model.

Only the top 20 variables and interactions are shown. Interactions between variables are indicated by the multiplication sign. The top eight unique variables (highlighted in bold) by average rank were used in the final model including their interactions with age.

| **Variable** | **Average Rank** | **Times Selected in Top 8** |
| --- | --- | --- |
| **MUAC** | **1·0** | **10** |
| **Weight for age z-score** | **2·5** | **10** |
| **SpO_2_** | **2·8** | **10** |
| **How long since last admission, 7 days – 1 month** | **5·8** | **10** |
| **HIV+** | **5·9** | **8** |
| **Age × Water source, bore hole** | **6·6** | **7** |
| **Boil/disinfect/filter water** | **7·3** | **6** |
| Temperature | 8·3 | 4 |
| Abnormal BCS | 9·0 | 6 |
| Temperature-squared | 10·1 | 4 |
| Age × Time it took to reach hospital, >1 hour | 11·7 | 3 |
| Water source, municipal water | 12·5 | 2 |
| How long since last admission, <7 days | 15·3 | 0 |
| How long since last admission, >1 year | 15·4 | 0 |
| Age × How long since last admission, 1 month – 1 year | 16·5 | 0 |
| Respiratory rate | 16·6 | 0 |
| Age × Water source, municipal water | 17·3 | 0 |
| Malaria | 18·4 | 0 |
| Time it took to reach hospital, >1 hour | 18·4 | 0 |
| Maternal education | 19·0 | 0 |

Abbreviations: HIV = human immunodeficiency virus; MUAC = mid-upper arm circumference; SpO_2_ = oxygen saturation

### S5: Intermediary Any Variable Models – Performance Metrics, Coefficients and Variable Importance

#### **Table S5.1** Performance metrics across 10 folds of cross-validation from the intermediary **0-6-month** any variable model using the probability threshold that gave 80% sensitivity

| **Fold** | **Specificity** | **Sensitivity** | **AUC** | **PPV** | **NPV** | **PRAUC** | **Brier Score** |
| --- | --- | --- | --- | --- | --- | --- | --- |
| 1 | 0·748 | 0·808 | 0·847 | 0·212 | 0·979 | 0·307 | 0·063 |
| 2 | 0·518 | 0·808 | 0·727 | 0·124 | 0·970 | 0·165 | 0·069 |
| 3 | 0·539 | 0·808 | 0·701 | 0·128 | 0·971 | 0·209 | 0·068 |
| 4 | 0·825 | 0·800 | 0·869 | 0·270 | 0·981 | 0·320 | 0·060 |
| 5 | 0·455 | 0·808 | 0·699 | 0·111 | 0·966 | 0·231 | 0·067 |
| 6 | 0·605 | 0·800 | 0·768 | 0·141 | 0·974 | 0·193 | 0·065 |
| 7 | 0·738 | 0·808 | 0·803 | 0·206 | 0·979 | 0·195 | 0·068 |
| 8 | 0·621 | 0·808 | 0·782 | 0·152 | 0·975 | 0·200 | 0·069 |
| 9 | 0·667 | 0·800 | 0·821 | 0·163 | 0·976 | 0·203 | 0·065 |
| 10 | 0·521 | 0·808 | 0·691 | 0·124 | 0·970 | 0·128 | 0·072 |
| **Average** | **0·624** | **0·805** | **0·771** | **0·163** | **0·974** | **0·215** | **0·067** |

Abbreviations: AUC = area under the receiver operating characteristic curve; PPV = positive predictive value; NPV = negative predictive value; PRAUC = area under the precision-recall curve

#### **Table S5.2** Performance metrics across 10 folds of cross-validation from the intermediary **6-60-month** any variable model using the probability threshold that gave 80% sensitivity

| **Fold** | **Specificity** | **Sensitivity** | **AUC** | **PPV** | **NPV** | **PRAUC** | **Brier Score** |
| --- | --- | --- | --- | --- | --- | --- | --- |
| 1 | 0·652 | 0·783 | 0·751 | 0·101 | 0·984 | 0·140 | 0·044 |
| 2 | 0·481 | 0·792 | 0·720 | 0·074 | 0·978 | 0·162 | 0·045 |
| 3 | 0·509 | 0·783 | 0·707 | 0·074 | 0·979 | 0·111 | 0·045 |
| 4 | 0·557 | 0·783 | 0·719 | 0·081 | 0·981 | 0·141 | 0·044 |
| 5 | 0·680 | 0·783 | 0·773 | 0·109 | 0·984 | 0·206 | 0·042 |
| 6 | 0·553 | 0·783 | 0·747 | 0·081 | 0·981 | 0·143 | 0·043 |
| 7 | 0·554 | 0·783 | 0·793 | 0·081 | 0·981 | 0·181 | 0·043 |
| 8 | 0·560 | 0·783 | 0·753 | 0·082 | 0·981 | 0·182 | 0·043 |
| 9 | 0·724 | 0·792 | 0·812 | 0·130 | 0·985 | 0·177 | 0·045 |
| 10 | 0·728 | 0·792 | 0·814 | 0·132 | 0·985 | 0·186 | 0·044 |
| **Average** | **0·600** | **0·785** | **0·759** | **0·094** | **0·982** | **0·163** | **0·044** |

Abbreviations: AUC = area under the receiver operating characteristic curve; PPV = positive predictive value; NPV = negative predictive value; PRAUC = area under the precision-recall curve

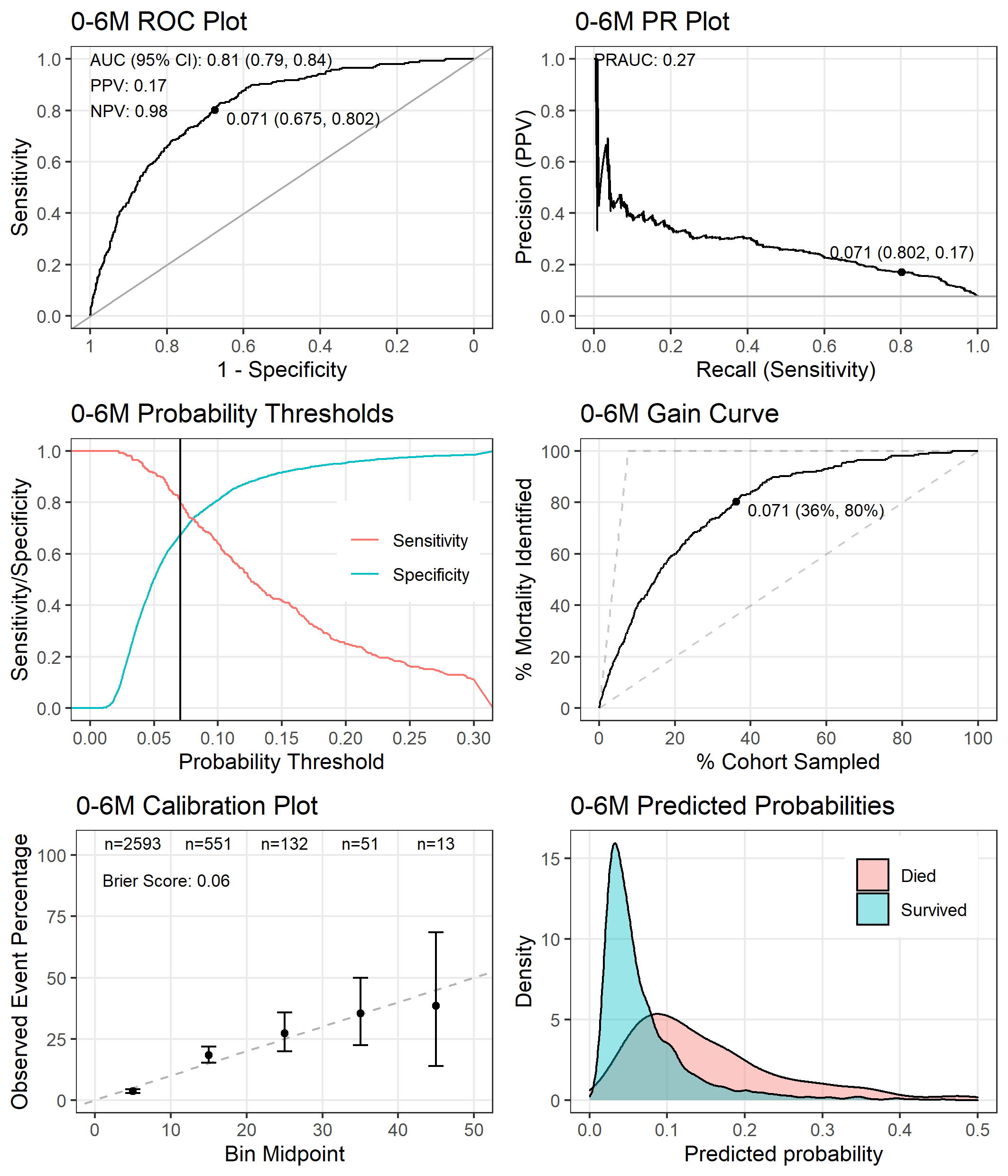

#### **Figure S5.1** Performance of the intermediary **0-6-month** any variable model tested on the entire dataset.

The point on the receiver operating characteristic (ROC) plot, precision recall (PR) plot, and gain curve indicates the co-ordinates when using the probability threshold that gives a sensitivity of 80% (probability threshold = 0·071). The positive predictive value (PPV) and negative predictive value (NPV) are also reported in the ROC plot using this threshold.

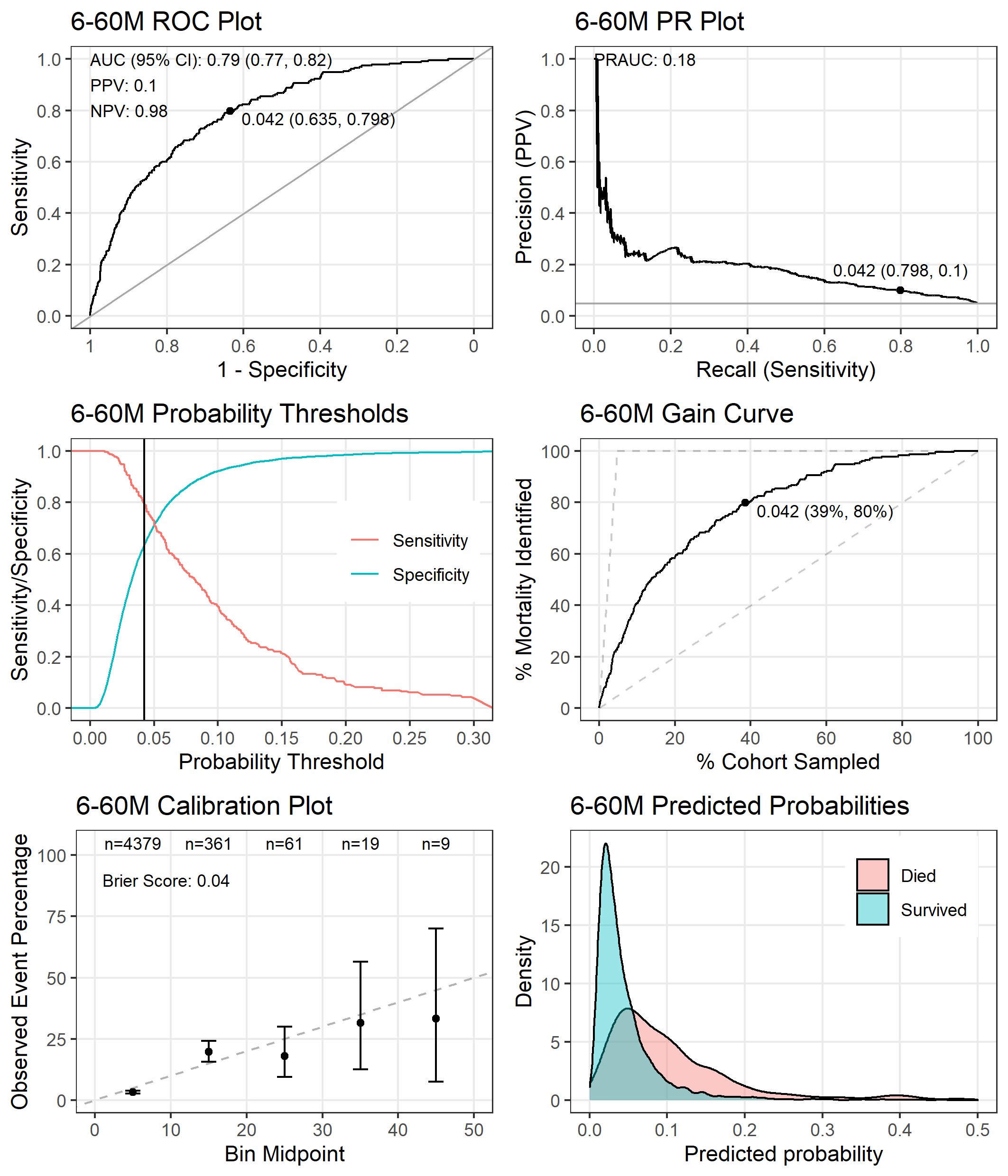

#### **Figure S5.2** Performance of the intermediary **6-60-month** any variable model tested on the entire dataset.

The point on the receiver operating characteristic (ROC) plot, precision recall (PR) plot, and gain curve indicates the co-ordinates when using the probability threshold that gives a sensitivity of 80% (probability threshold = 0·042). The positive predictive value (PPV) and negative predictive value (NPV) are also reported in the ROC plot using this threshold.

#### **Table S5.3** Coefficients of the intermediary **0-6-month** any variable model

| **Variable** | **Coefficient** |
| --- | --- |
| Intercept | -2·771 |
| Abdominal distension | 0·045 |
| Abnormal BCS | 0·022 |
| Number of children | -0·001 |
| BMI z-score | -0·009 |
| Dehydration WHO categories, severe dehydration | 0·013 |
| Duration of present illness, 8 days – 1 month | 0·094 |
| Duration of present illness, >1 month | 0·028 |
| Fontanelle | 0·087 |
| Glucose | 0·042 |
| Heart rate | -0·003 |
| Referral visit | 0·034 |
| Lactate | 0·065 |
| Malaria | -0·078 |
| Mother currently acutely ill | -0·029 |
| Maternal age | 0·009 |
| Maternal education | -0·069 |
| Maternal HIV, yes | 0·007 |
| MUAC | -0·257 |
| Neonate | -0·078 |
| Pallor | 0·047 |
| Prior care sought for current illness | 0·064 |
| How long since last admission, 7 days – 1 month | 0·049 |
| How long since last admission, 1 month – 1 year | 0·053 |
| Sex, male | 0·009 |
| SpO_2_ | -0·097 |
| Sucking well when breastfeeding, or feeding well if not breastfed | -0·135 |
| Temperature | -0·018 |
| Temperature-squared | -0·018 |
| When did the baby cry after birth, immediately | 0·063 |
| When did the baby cry after birth, 11-30 minutes | 0·009 |
| Abnormal tone | 0·075 |
| Time to reach hospital, 30 minutes – 1 hour | -0·036 |
| Time to reach hospital, >1 hour | 0·146 |
| Decreased urine production | 0·051 |
| Water source, municipal water | -0·025 |
| Water source, open source | 0·051 |
| Water source, fast running water | -0·032 |
| Weight for age z-score | -0·255 |
| Age × Abdominal distension | 0·012 |
| Age × Number of children | -0·045 |
| Age × Bed net use, sometimes | -0·020 |
| Age × Dehydration WHO categories, some dehydration | 0·021 |
| Age × Dehydration WHO categories, severe dehydration | 0·025 |
| Age × Delivery method, caesarean | -0·010 |
| Age × Diastolic blood pressure | -0·002 |
| Age × Glucose | 0·014 |
| Age × Referral visit | -0·010 |
| Age × Neonatal jaundice | 0·092 |
| Age × Malaria | -0·033 |
| Age × Mother currently acutely ill | -0·029 |
| Age × Maternal HIV, unknown | -0·017 |
| Age × Neonate | 0 |
| Age × Pallor | 0·027 |
| Age × Premature birth | -0·001 |
| Age × How long since last admission, <7 days | 0·073 |
| Age × How long since last admission, 7 days – 1 month | 0·011 |
| Age × SpO_2_ transformed | 0·034 |
| Age × Sucking well when breastfeeding, or feeding well if not breastfed, prior to illness | 0·058 |
| Age × Systolic blood pressure | -0·002 |
| Age × When did the baby cry after birth, immediately | 0·024 |
| Age × When did the baby cry after birth, <5 minutes | 0·009 |
| Age × Abnormal tone | 0·016 |
| Age × Boil/disinfect/filter water | 0·047 |
| Age × Weight for age z-score | -0·050 |

Interactions between variables are indicated by the multiplication sign.

Abbreviations: BCS = Blantyre coma scale; BMI = body mass index; HIV = human immunodeficiency virus; MUAC = mid-upper arm circumference; SpO_2_ = oxygen saturation; WHO = World Health Organisation

#### **Table S5.4** Coefficients of the intermediary **6-60-month** any variable model.

| **Variable** | **Coefficient** |
| --- | --- |
| Intercept | -3·297 |
| Abnormal BCS | 0·099 |
| Bed net use, sometimes | 0·008 |
| Haemoglobin | -0·223 |
| HIV+ | 0·112 |
| Malaria | -0·113 |
| Maternal education | -0·029 |
| Maternal HIV, unknown | 0·019 |
| MUAC | -0·378 |
| How long since last admission, <7 days | 0·063 |
| How long since last admission, 7 days – 1 month | 0·128 |
| How long since last admission, 1 month – 1 year | 0·023 |
| How long since last admission, >1 year | -0·058 |
| Respiratory rate | 0·023 |
| SpO_2_ | -0·162 |
| Temperature | -0·104 |
| Temperature-squared | -0·096 |
| Time it took to reach hospital, >1 hour | 0·006 |
| Boil/disinfect/filter water | -0·095 |
| Water source, municipal water | -0·061 |
| Weight for age z-score | -0·223 |
| Age × Abnormal BCS | -0·008 |
| Age × Diastolic blood pressure | 0·001 |
| Age × How long since last admission, 7 days – 1 month | 0·005 |
| Age × How long since last admission, 1 month – 1 year | 0·060 |
| Age × Respiratory rate | 0·045 |
| Age × Time it took to reach hospital, >1 hour | 0·088 |
| Age × Water source, bore hole | 0·130 |
| Age × Water source, municipal water | -0·030 |
| Age × Weight for age z-score | 0·130 |

Interactions between variables are indicated by the multiplication sign.

Abbreviations: BCS = Blantyre coma scale; HIV = human immunodeficiency virus; MUAC = mid-upper arm circumference; SpO_2_ = oxygen saturation

#### **Table S5.5** Average rank of variable importance and the number of times selected in the top 8 variables across 10 folds of cross-validation from the intermediary **0-6-month** any variable model.

Only the top 20 variables and interactions are shown. Interactions between variables are indicated by the multiplication sign. The top eight unique variables by average rank are highlighted in bold. Note that top eight variables are identical to those selected by the intermediary clinical and social model (see Table S4.1).

| **Variable** | **Average Rank** | **Times Selected in Top 8** |
| --- | --- | --- |
| **Weight for age z-score** | **1·1** | **10** |
| **MUAC** | **1·9** | **10** |
| **Time it took to reach hospital, >1 hour** | **3·2** | **10** |
| **Sucking well when breastfeeding, or feeding well if not breastfed** | **4·2** | **10** |
| **SpO_2_** | **7**·**0** | **7** |
| **Duration of present illness, 8 days – 1 month** | **7·9** | **7** |
| **Age × Neonatal jaundice** | **8·3** | **6** |
| Abnormal tone | 10·3 | 4 |
| Fontanelle | 11·2 | 7 |
| Malaria | 14·0 | 1 |
| Neonate | 15·1 | 2 |
| Age × How long since last admission, <7 days | 15·1 | 1 |
| Prior care sought for current illness | 16·4 | 0 |
| Lactate | 16·7 | 0 |
| Maternal education | 17·6 | 1 |
| When did the baby cry after birth, immediately | 18·4 | 0 |
| Water source, open source | 20·8 | 0 |
| How long since last admission, 1 month – 1 year | 22·0 | 0 |
| Decreased urine production | 22·1 | 0 |
| How long since last admission, 7 days – 1 month | 24·7 | 0 |

Abbreviations: MUAC = mid-upper arm circumference; SpO_2_ = oxygen saturation

#### **Table S5.6** Average rank of variable importance and the number of times selected in the top 8 variables across 10 folds of cross-validation from the intermediary **6-60-month** any variable model.

Only the top 20 variables and interactions are shown. Interactions between variables are indicated by the multiplication sign. The top eight unique variables by average rank are highlighted in bold.

| **Variable** | **Average Rank** | **Times Selected in Top 8** |
| --- | --- | --- |
| **MUAC** | **1·0** | **10** |
| **Haemoglobin** | **2·5** | **10** |
| **Weight for age z-score** | **2·7** | **10** |
| **SpO_2_** | **4·5** | **9** |
| **How long since last admission, 7 days – 1 month** | **7·1** | **7** |
| **Age × Water source, bore hole** | **7·9** | **5** |
| **HIV+** | **8·2** | **7** |
| Age × Weight for length z-score | 8·3 | 6 |
| Temperature | 9·7 | 2 |
| Malaria | 10·5 | 4 |
| Abnormal BCS | 11·2 | 3 |
| Boil/disinfect/filter water | 11·5 | 3 |
| Temperature-squared | 11·7 | 1 |
| Age × Time to reach hospital, >1 hour | 12·6 | 3 |
| Water source, municipal water | 15·8 | 0 |
| How long since last admission, <7 days | 16·5 | 0 |
| How long since last admission, >1 year | 17·8 | 0 |
| Age × How long since last admission, 1 month – 1 year | 15·9 | 0 |
| Age × Respiratory rate | 22·2 | 0 |
| Maternal education | 24·1 | 0 |

Abbreviations: BCS = Blantyre coma scale; HIV = human immunodeficiency virus; MUAC = mid-upper arm circumference; SpO_2_ = oxygen saturation

### S6: Final Clinical Variable Models, M6PD-C_0-6_ and M6PD-C_6-60_ – Performance Metrics and Coefficients

#### **Table S6.1** Performance metrics across 10 folds of cross-validation from the **M6PD-C_0-6_ model** using the probability threshold that gave 80% sensitivity.

The top eight unique variables with the highest average variable importance from 10-fold cross-validation in the intermediary clinical variable model were used (see Table S3.1).

| **Fold** | **Specificity** | **Sensitivity** | **AUC** | **PPV** | **NPV** | **PRAUC** | **Brier Score** |
| --- | --- | --- | --- | --- | --- | --- | --- |
| 1 | 0·757 | 0·808 | 0·848 | 0·219 | 0·979 | 0·331 | 0·064 |
| 2 | 0·440 | 0·808 | 0·688 | 0·108 | 0·965 | 0·159 | 0·070 |
| 3 | 0·616 | 0·808 | 0·722 | 0·150 | 0·974 | 0·186 | 0·068 |
| 4 | 0·725 | 0·800 | 0·814 | 0·190 | 0·978 | 0·302 | 0·063 |
| 5 | 0·429 | 0·808 | 0·694 | 0·106 | 0·964 | 0·248 | 0·067 |
| 6 | 0·589 | 0·800 | 0·763 | 0·136 | 0·973 | 0·296 | 0·063 |
| 7 | 0·663 | 0·808 | 0·795 | 0·168 | 0·976 | 0·196 | 0·068 |
| 8 | 0·715 | 0·808 | 0·795 | 0·193 | 0·978 | 0·206 | 0·068 |
| 9 | 0·583 | 0·800 | 0·794 | 0·134 | 0·973 | 0·218 | 0·065 |
| 10 | 0·469 | 0·808 | 0·632 | 0·114 | 0·967 | 0·113 | 0·072 |
| **Average** | **0·599** | **0·805** | **0·754** | **0·152** | **0·973** | **0·226** | **0·067** |

Abbreviations: AUC = area under the receiver operating characteristic curve; PPV = positive predictive value; NPV = negative predictive value; PRAUC = area under the precision-recall curve

#### **Table S6.2** Performance metrics across 10 folds of cross-validation from the **M6PD-C_6-60_ model** using the probability threshold that gave 80% sensitivity.

The top nine unique variables with the highest average variable importance from 10-fold cross-validation in the intermediary clinical variable model were used (see Table S3.2).

| **Fold** | **Specificity** | **Sensitivity** | **AUC** | **PPV** | **NPV** | **PRAUC** | **Brier Score** |
| --- | --- | --- | --- | --- | --- | --- | --- |
| 1 | 0·737 | 0·783 | 0·768 | 0·129 | 0·985 | 0·139 | 0·043 |
| 2 | 0·606 | 0·792 | 0·748 | 0·095 | 0·982 | 0·171 | 0·045 |
| 3 | 0·483 | 0·783 | 0·675 | 0·070 | 0·978 | 0·099 | 0·045 |
| 4 | 0·304 | 0·783 | 0·665 | 0·053 | 0·966 | 0·139 | 0·043 |
| 5 | 0·535 | 0·783 | 0·750 | 0·078 | 0·980 | 0·214 | 0·042 |
| 6 | 0·588 | 0·783 | 0·756 | 0·087 | 0·982 | 0·173 | 0·042 |
| 7 | 0·580 | 0·783 | 0·737 | 0·085 | 0·982 | 0·137 | 0·044 |
| 8 | 0·549 | 0·783 | 0·699 | 0·080 | 0·981 | 0·149 | 0·044 |
| 9 | 0·772 | 0·792 | 0·798 | 0·153 | 0·986 | 0·166 | 0·045 |
| 10 | 0·520 | 0·792 | 0·723 | 0·079 | 0·98 | 0·187 | 0·044 |
| **Average** | **0·567** | **0·785** | **0·732** | **0·091** | **0·98** | **0·157** | **0·044** |

Abbreviations: AUC = area under the receiver operating characteristic curve; PPV = positive predictive value; NPV = negative predictive value; PRAUC = area under the precision-recall curve

#### **Table S6.3** Coefficients of the **M6PD-C_0-6_ model**.

| **Variable** | **Coefficient** |
| --- | --- |
| Intercept | -2·760 |
| Weight for age z-score | -0·344 |
| MUAC | -0·295 |
| Sucking well when breastfeeding, or feeding well if not breastfed | -0·248 |
| SpO_2_ | -0·165 |
| Duration of present illness, 48 hours – 7 days | 0·023 |
| Duration of present illness, 8 days – 1 month | 0·194 |
| Duration of present illness, >1 month | 0·068 |
| Age | 0·068 |
| Fontanelle | 0·134 |
| Age × Weight for age z-score | -0·054 |
| Age × Sucking well when breastfeeding | 0·023 |
| Age × Duration of present illness, 48 hours – 7 days | 0·060 |
| Age × Jaundice | 0·101 |

Interactions between variables are indicated by the multiplication sign.

Abbreviations: MUAC = mid-upper arm circumference; SpO_2_ = oxygen saturation

#### **Table S6.4** Coefficients of the **M6PD-C_6-60_ model**.

| **Variable** | **Coefficient** |
| --- | --- |
| Intercept | -3·243 |
| MUAC | -0·411 |
| SpO_2_ | -0·186 |
| Weight for age z-score | -0·177 |
| Age | 0·029 |
| How long since last admission, <7 days | 0·083 |
| How long since last admission, 7 days – 1 month | 0·142 |
| How long since last admission, 1 month – 1 year | 0·030 |
| How long since last admission, >1 year | -0·081 |
| Respiratory rate | 0·054 |
| Abnormal BCS | 0·156 |
| Temperature, °C | -0·123 |
| Temperature-squared, °C | -0·116 |
| HIV+ | 0·127 |
| Age × MUAC | 0·016 |
| Age × SpO_2_ | 0·011 |
| Age × How long since last admission, <7 days | 0·026 |
| Age × How long since last admission, 7 days – 1 month | 0·023 |
| Age × How long since last admission, 1 month – 1 year | 0·086 |
| Age × How long since last admission >1 year | -0·012 |
| Age × Respiratory rate | 0·106 |
| Age × Abnormal BCS | -0·050 |
| Age × HIV+ | -0·021 |

Interactions between variables are indicated by the multiplication sign.

Abbreviations: BCS = Blantyre coma scale; HIV = human immunodeficiency virus; MUAC = mid-upper arm circumference; SpO_2_ = oxygen saturation

#### **Figure S6.1** Variable importance plot of the **M6PD-C_0-6_** model

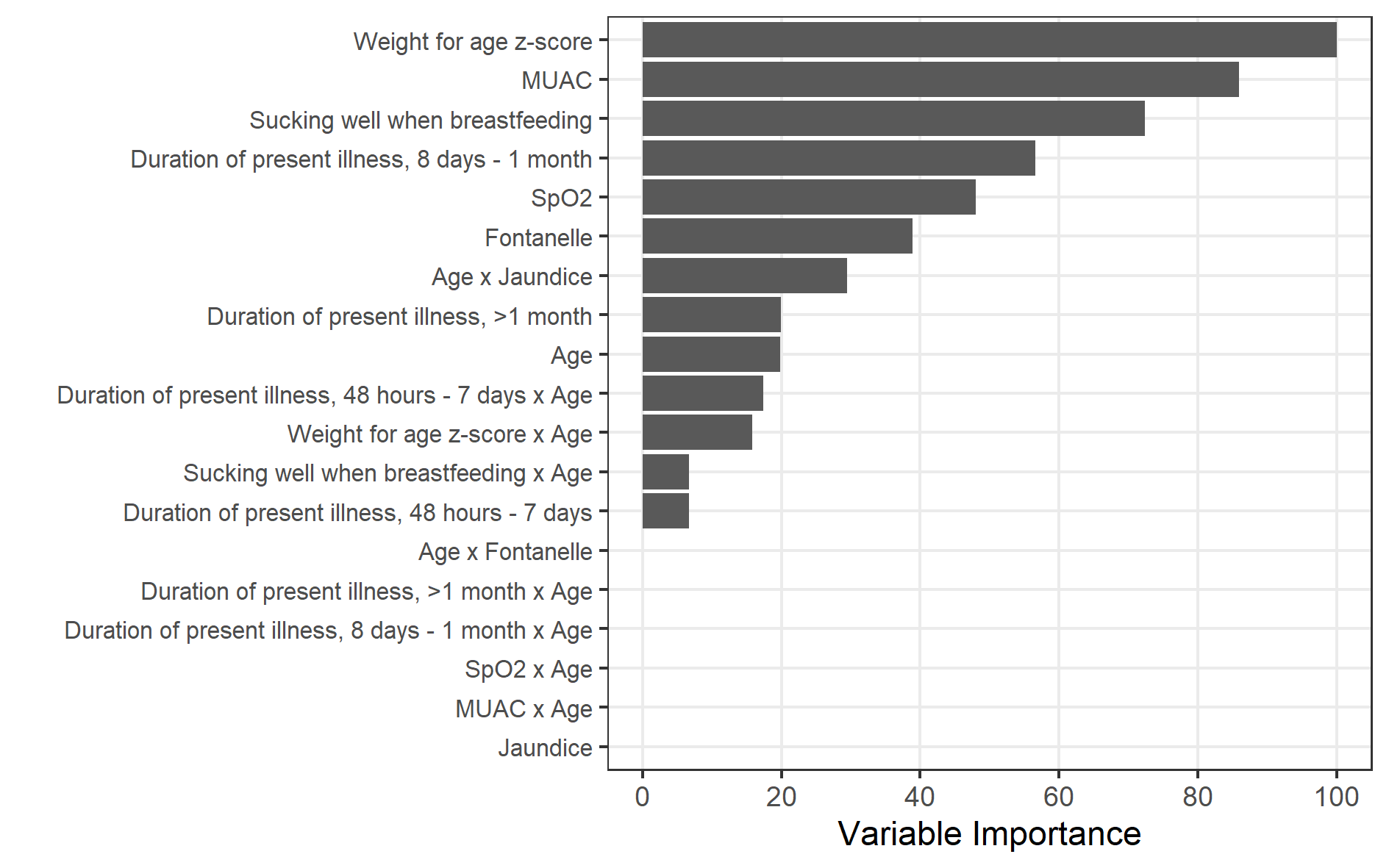

#### **Figure S6.2** Variable importance plot of the **M6PD-C_6-60_** model

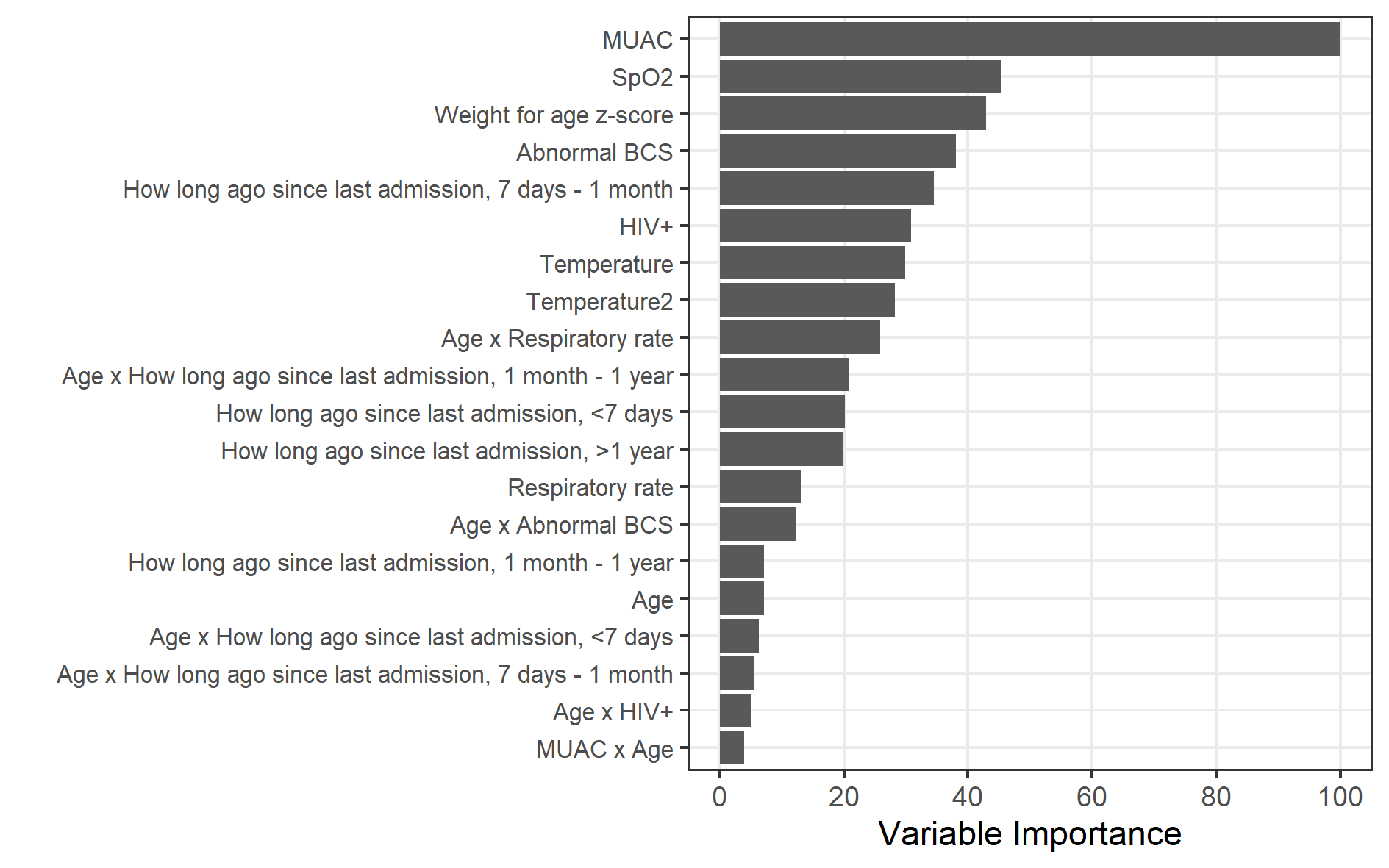

### S7: Final Clinical and Social Variable Models, M6PD-CS_0-6_ and M6PD-CS_6-60_ – Performance Metrics and Coefficients

Note that the **M6PD-CS_0-6_ model** is identical to the **M6PD-A_0-6_ model** reported in Supplementary Material S8.

#### **Table S7.1** Performance metrics across 10 folds of cross-validation from **M6PD-CS_0-6_ model** using the probability threshold that gave 80% sensitivity.

The top eight unique variables with the highest average variable importance from 10-fold cross-validation in the intermediary clinical and social variable model were used (see Table S4.1).

| **Fold** | **Specificity** | **Sensitivity** | **AUC** | **PPV** | **NPV** | **PRAUC** | **Brier Score** |
| --- | --- | --- | --- | --- | --- | --- | --- |
| 1 | 0·832 | 0·808 | 0·853 | 0·288 | 0·981 | 0·354 | 0·063 |
| 2 | 0·408 | 0·808 | 0·709 | 0·103 | 0·962 | 0·162 | 0·070 |
| 3 | 0·545 | 0·808 | 0·715 | 0·130 | 0·971 | 0·153 | 0·070 |
| 4 | 0·731 | 0·800 | 0·848 | 0·194 | 0·978 | 0·342 | 0·062 |
| 5 | 0·513 | 0·808 | 0·710 | 0·122 | 0·970 | 0·227 | 0·067 |
| 6 | 0·508 | 0·800 | 0·756 | 0·116 | 0·969 | 0·278 | 0·064 |
| 7 | 0·709 | 0·808 | 0·812 | 0·189 | 0·978 | 0·207 | 0·067 |
| 8 | 0·728 | 0·808 | 0·805 | 0·200 | 0·978 | 0·208 | 0·067 |
| 9 | 0·686 | 0·800 | 0·791 | 0·171 | 0·977 | 0·192 | 0·065 |
| 10 | 0·421 | 0·808 | 0·655 | 0·105 | 0·963 | 0·126 | 0·071 |
| **Average** | **0·608** | **0·805** | **0·765** | **0·162** | **0·973** | **0·225** | **0·067** |

Abbreviations: AUC = area under the receiver operating characteristic curve; PPV = positive predictive value; NPV = negative predictive value; PRAUC = area under the precision-recall curve

#### **Table S7.2** Performance metrics across 10 folds of cross-validation from **M6PD-CS_6-60_ model** using the probability threshold that gave 80% sensitivity.

The top eight unique variables with the highest average variable importance from 10-fold cross-validation in the intermediary clinical and social variable model were used (see Table S4.2).

| **Fold** | **Specificity** | **Sensitivity** | **AUC** | **PPV** | **NPV** | **PRAUC** | **Brier Score** |
| --- | --- | --- | --- | --- | --- | --- | --- |
| 1 | 0·570 | 0·783 | 0·729 | 0·083 | 0·981 | 0·108 | 0·044 |
| 2 | 0·473 | 0·792 | 0·681 | 0·073 | 0·977 | 0·118 | 0·046 |
| 3 | 0·702 | 0·783 | 0·769 | 0·116 | 0·985 | 0·129 | 0·044 |
| 4 | 0·441 | 0·783 | 0·652 | 0·065 | 0·976 | 0·131 | 0·044 |
| 5 | 0·683 | 0·783 | 0·766 | 0·110 | 0·984 | 0·210 | 0·042 |
| 6 | 0·651 | 0·783 | 0·761 | 0·101 | 0·984 | 0·169 | 0·043 |
| 7 | 0·622 | 0·783 | 0·762 | 0·094 | 0·983 | 0·154 | 0·043 |
| 8 | 0·488 | 0·783 | 0·713 | 0·071 | 0·978 | 0·171 | 0·043 |
| 9 | 0·628 | 0·792 | 0·784 | 0·100 | 0·983 | 0·137 | 0·045 |
| 10 | 0·674 | 0·792 | 0·793 | 0·112 | 0·984 | 0·210 | 0·044 |
| **Average** | **0·593** | **0·785** | **0·741** | **0·093** | **0·982** | **0·154** | **0·044** |

Abbreviations: AUC = area under the receiver operating characteristic curve; PPV = positive predictive value; NPV = negative predictive value; PRAUC = area under the precision-recall curve

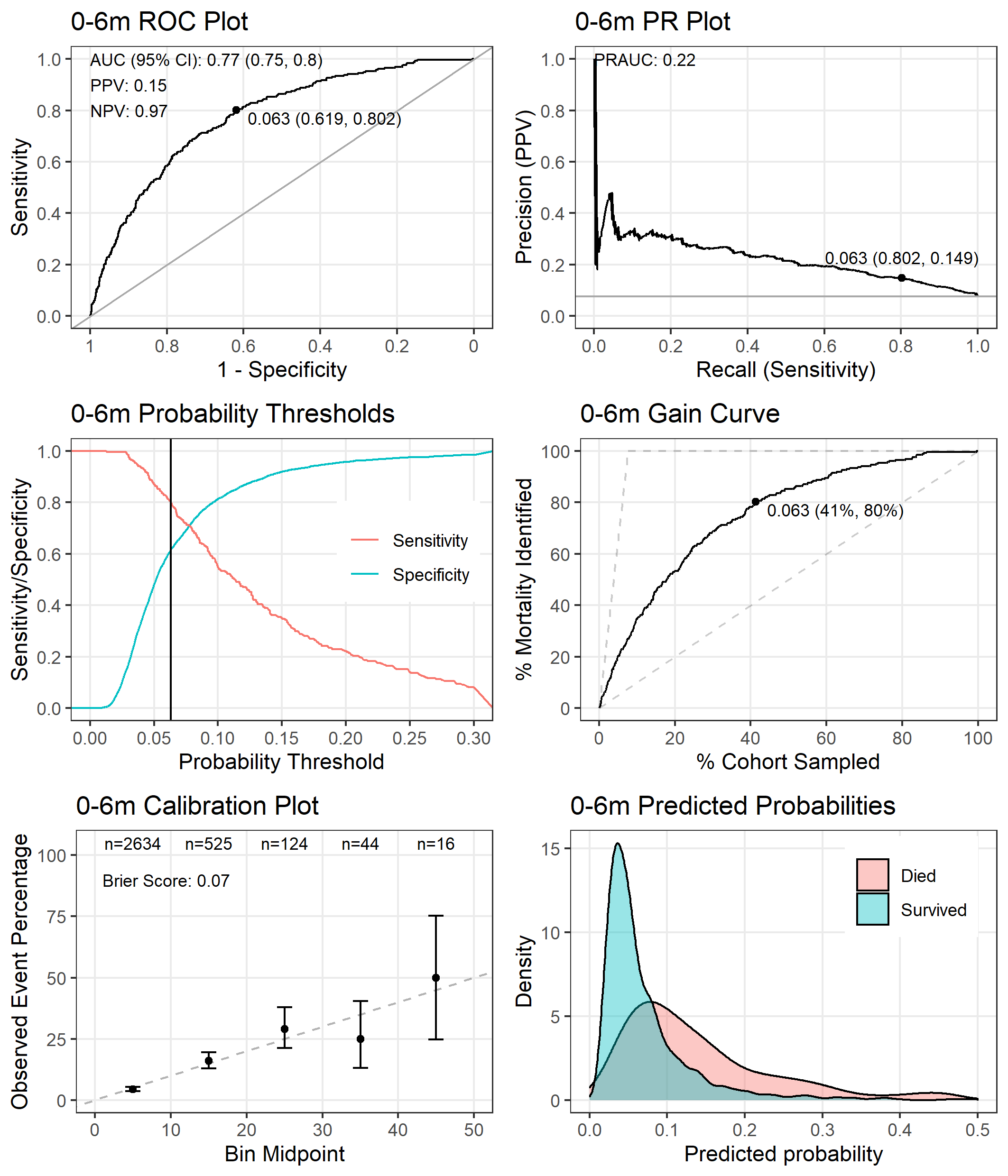

#### **Figure S7.1** Performance of the **M6PD-CS_0-6_ model** tested on the entire dataset.

The point on the receiver operating characteristic (ROC) plot, precision recall (PR) plot, and gain curve indicates the co-ordinates when using the probability threshold that gives a sensitivity of 80% (probability threshold = 0·063). The positive predictive value (PPV) and negative predictive value (NPV) are also reported in the ROC plot using this threshold.

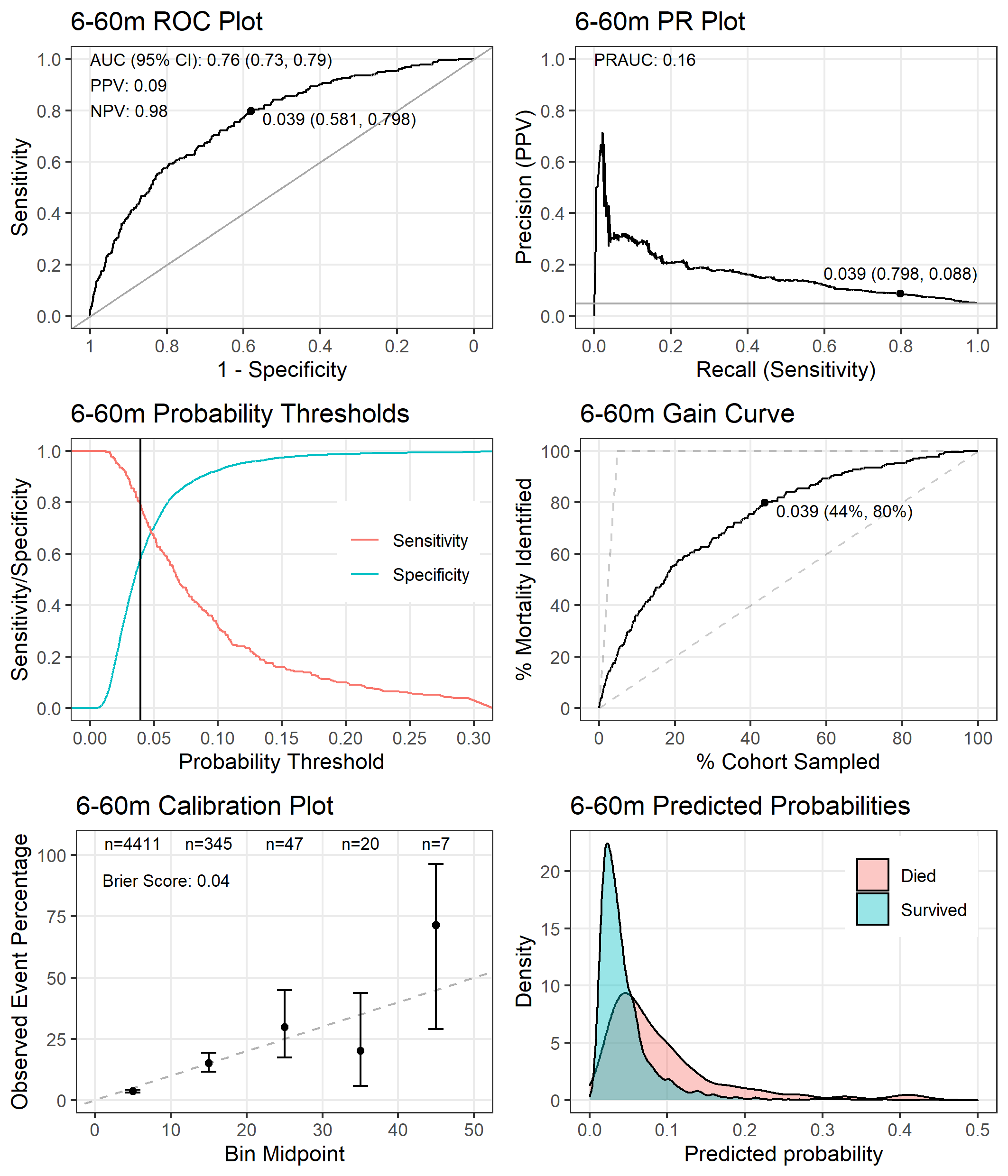

#### **Figure S7.2** Performance of the **M6PD-CS_6-60_ model** tested on the entire dataset.

The point on the receiver operating characteristic (ROC) plot, precision recall (PR) plot, and gain curve indicates the co-ordinates when using the probability threshold that gives a sensitivity of 80% (probability threshold = 0·039). The positive predictive value (PPV) and negative predictive value (NPV) are also reported in the ROC plot using this threshold.

#### **Table S7.3** Coefficients of the **M6PD-CS_0-6_ model**.

| **Variable** | **Coefficient** |
| --- | --- |
| Intercept | -2·753 |
| Weight for age z-score | -0·361 |
| MUAC | -0·219 |
| Time it took to reach hospital, >1 hour | 0·233 |
| Sucking well when breastfeeding, or feeding well if not breastfed | -0·206 |
| SpO_2_ | -0·149 |
| Duration of present illness, 8 days – 1 month | 0·130 |
| Duration of present illness, >1 month | 0·031 |
| Age | 0·031 |
| Age × Weight for age z-score | -0·064 |
| Age × Duration of present illness, 48 hours – 7 days | 0·005 |
| Age × Jaundice | 0·087 |

Interactions between variables are indicated by the multiplication sign.

Abbreviations: MUAC = mid-upper arm circumference; SpO_2_ = oxygen saturation

#### **Table S7.4** Coefficients of the **M6PD-CS_6-60_ model**.

| **Variable** | **Coefficient** |
| --- | --- |
| Intercept | -3·241 |
| MUAC | -0·386 |
| Weight for age z-score | -0·180 |
| SpO_2_ | -0·186 |
| HIV+ | 0·111 |
| Age | 0·007 |
| Water source, municipal water | -0·102 |
| How long since last admission, <7 days | 0·074 |
| How long since last admission, 7 days – 1 month | 0·124 |
| How long since last admission, 1 month – 1 year | 0·013 |
| How long since last admission, >1 year | -0·059 |
| Boil/disinfect/filter water | -0·140 |
| Age × Water source, bore hole | 0·157 |
| Age × Water source, municipal water | -0·044 |
| Age × How long since last admission, <7 days | 0·006 |
| Age × How long since last admission, 7 days – 1 month | 0·007 |
| Age × How long since last admission, 1 month – 1 year | 0·077 |

Interactions between variables are indicated by the multiplication sign.

Abbreviations: HIV = human immunodeficiency virus; MUAC = mid-upper arm circumference; SpO_2_ = oxygen saturation

#### **Figure S7.3** Variable importance plot of the **M6PD-CS_0-6_** model

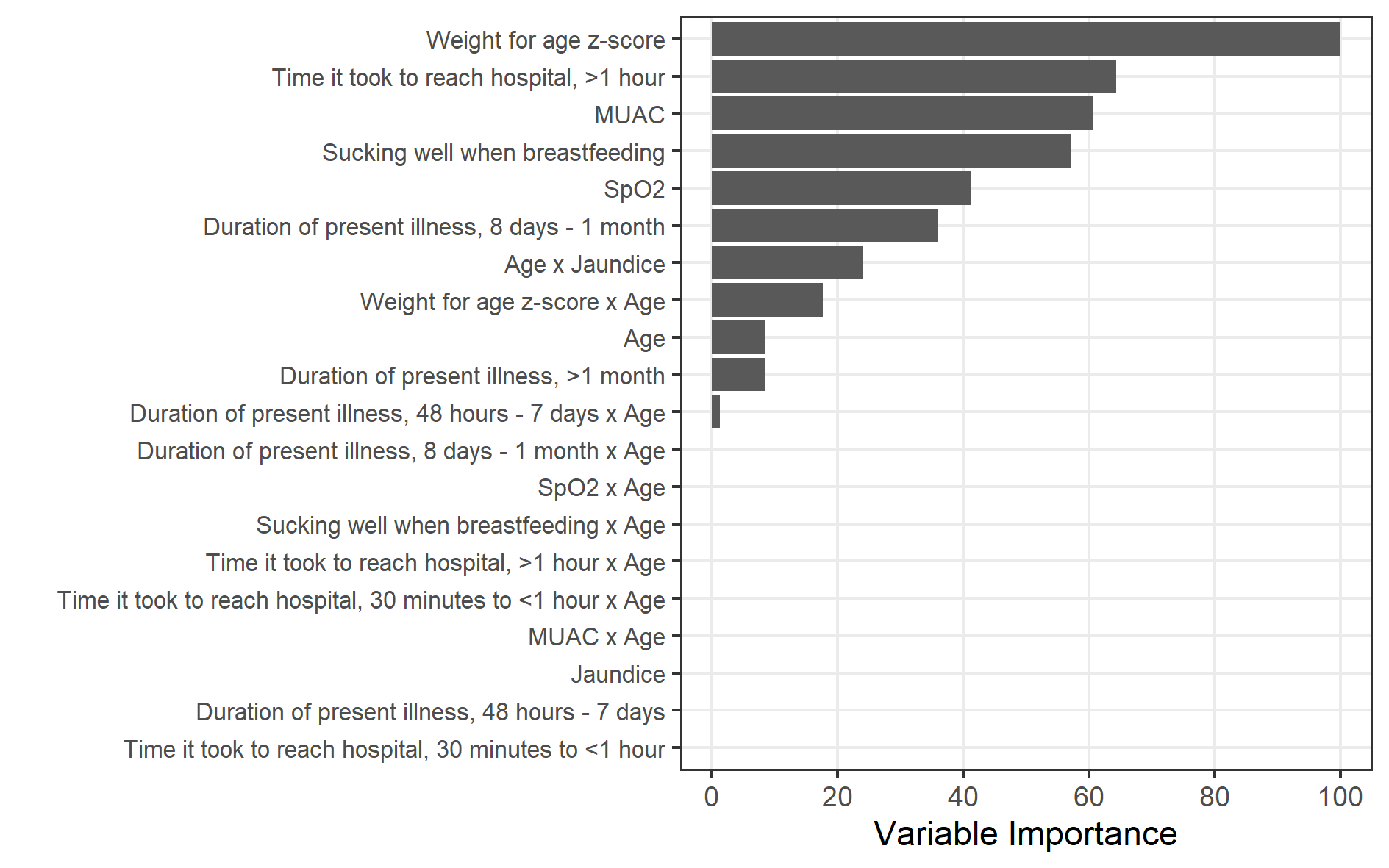

#### **Figure S7.4** Variable importance plot of the **M6PD-CS_6-60_** model

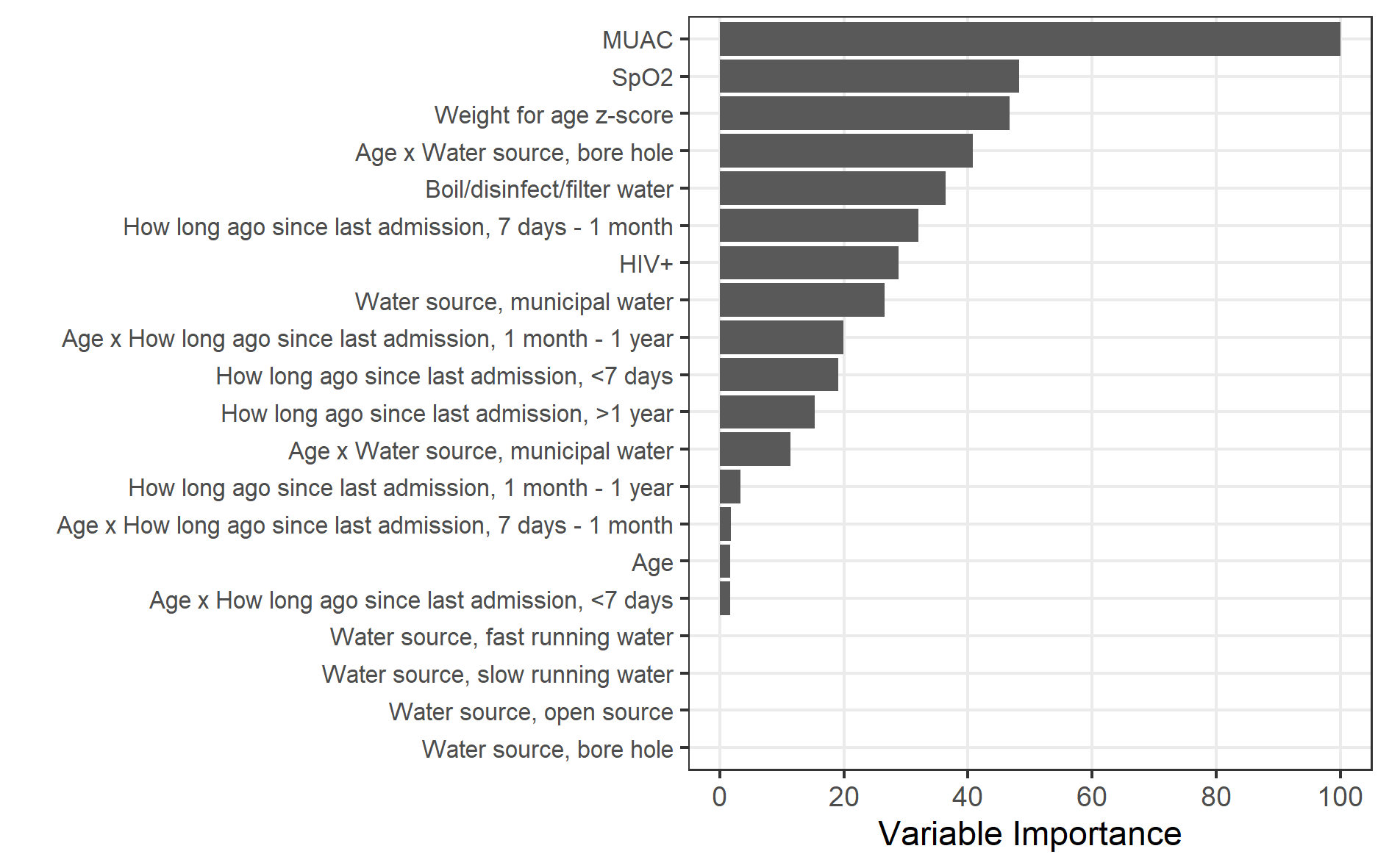

### S8: Final Any Variable Models, M6PD-A_0-6_ and M6PD-A_6-60_ – Performance Metrics and Coefficients

Note that **M6PD-A_0-6_ model** is identical to the **M6PD-CS_0-6_** model reported in Supplementary Material S7.

#### **Table S8.1** Performance metrics across 10 folds of cross-validation from **M6PD-A_0-6_ model** using the probability threshold that gave 80% sensitivity.

The top eight unique variables with the highest average variable importance from 10-fold cross-validation in the intermediary any variable model were used (see Table S5.5).

| **Fold** | **Specificity** | **Sensitivity** | **AUC** | **PPV** | **NPV** | **PRAUC** | **Brier Score** |
| --- | --- | --- | --- | --- | --- | --- | --- |
| 1 | 0·832 | 0·808 | 0·853 | 0·288 | 0·981 | 0·354 | 0·063 |
| 2 | 0·408 | 0·808 | 0·709 | 0·103 | 0·962 | 0·162 | 0·070 |
| 3 | 0·545 | 0·808 | 0·715 | 0·130 | 0·971 | 0·153 | 0·070 |
| 4 | 0·731 | 0·800 | 0·848 | 0·194 | 0·978 | 0·342 | 0·062 |
| 5 | 0·513 | 0·808 | 0·710 | 0·122 | 0·970 | 0·227 | 0·067 |
| 6 | 0·508 | 0·800 | 0·756 | 0·116 | 0·969 | 0·278 | 0·064 |
| 7 | 0·709 | 0·808 | 0·812 | 0·189 | 0·978 | 0·207 | 0·067 |
| 8 | 0·728 | 0·808 | 0·805 | 0·200 | 0·978 | 0·208 | 0·067 |
| 9 | 0·686 | 0·800 | 0·791 | 0·171 | 0·977 | 0·192 | 0·065 |
| 10 | 0·421 | 0·808 | 0·655 | 0·105 | 0·963 | 0·126 | 0·071 |
| **Average** | **0·608** | **0·805** | **0·765** | **0·162** | **0·973** | **0·225** | **0·067** |

Abbreviations: AUC = area under the receiver operating characteristic curve; PPV = positive predictive value; NPV = negative predictive value; PRAUC = area under the precision-recall curve

#### **Table S8.2** Performance metrics across 10 folds of cross-validation from the **M6PD-A_6-60_ model** using the probability threshold that gave 80% sensitivity.

The top eight unique variables with the highest average variable importance from 10-fold cross-validation in the intermediary any variable model were used (see Table S5·6).

| **Fold** | **Specificity** | **Sensitivity** | **AUC** | **PPV** | **NPV** | **PRAUC** | **Brier Score** |
| --- | --- | --- | --- | --- | --- | --- | --- |
| 1 | 0·511 | 0·783 | 0·712 | 0·074 | 0·979 | 0·110 | 0·044 |
| 2 | 0·420 | 0·792 | 0·693 | 0·067 | 0·975 | 0·121 | 0·046 |
| 3 | 0·424 | 0·783 | 0·703 | 0·064 | 0·975 | 0·103 | 0·045 |
| 4 | 0·502 | 0·783 | 0·690 | 0·073 | 0·979 | 0·121 | 0·044 |
| 5 | 0·513 | 0·783 | 0·747 | 0·074 | 0·979 | 0·196 | 0·043 |
| 6 | 0·516 | 0·783 | 0·760 | 0·075 | 0·979 | 0·153 | 0·043 |
| 7 | 0·563 | 0·783 | 0·778 | 0·082 | 0·981 | 0·151 | 0·043 |
| 8 | 0·477 | 0·783 | 0·735 | 0·070 | 0·978 | 0·180 | 0·043 |
| 9 | 0·715 | 0·792 | 0·811 | 0·127 | 0·985 | 0·158 | 0·045 |
| 10 | 0·709 | 0·792 | 0·830 | 0·124 | 0·985 | 0·214 | 0·044 |
| **Average** | **0·535** | **0·785** | **0·746** | **0·083** | **0·979** | **0·151** | **0·044** |

Abbreviations: AUC = area under the receiver operating characteristic curve; PPV = positive predictive value; NPV = negative predictive value; PRAUC = area under the precision-recall curve

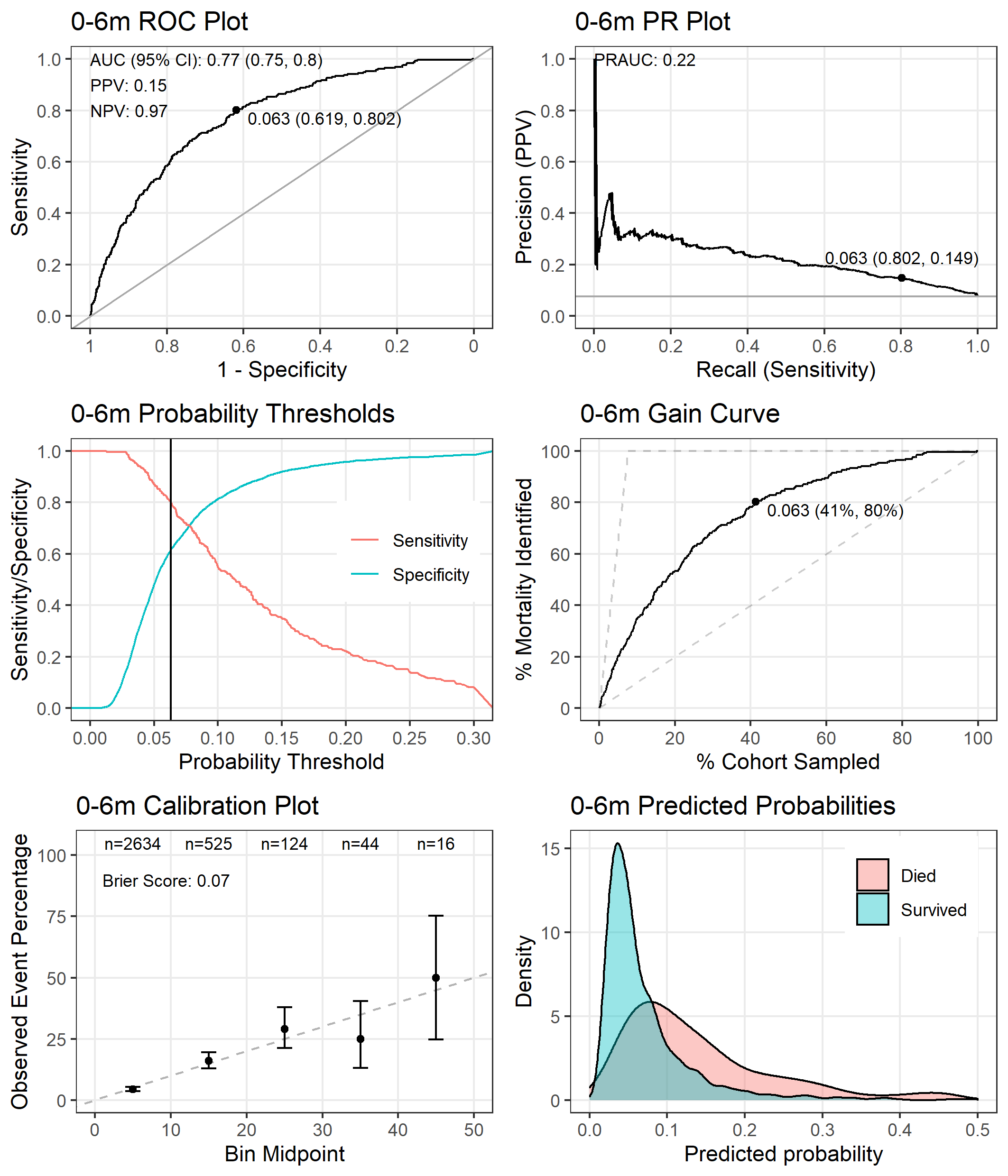

#### **Figure S8.1** Performance of the **M6PD-A_0-6_ model** tested on the entire dataset.

The point on the receiver operating characteristic (ROC) plot, precision recall (PR) plot, and gain curve indicates the co-ordinates when using the probability threshold that gives a sensitivity of 80% (probability threshold = 0·063). The positive predictive value (PPV) and negative predictive value (NPV) are also reported in the ROC plot using this threshold.

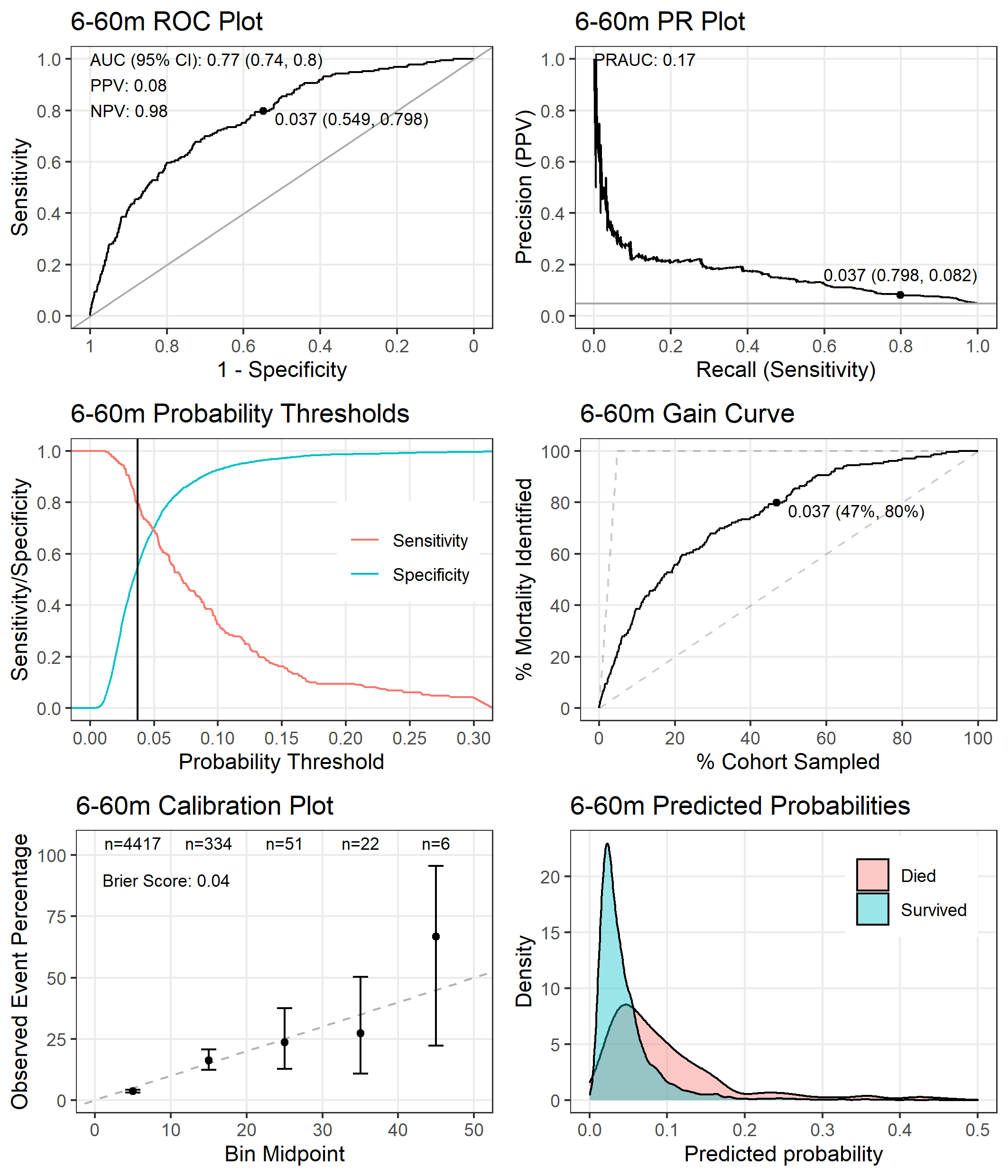

#### **Figure S8.2** Performance of the **M6PD-A_6-60_ model** tested on the entire dataset.

The point on the receiver operating characteristic (ROC) plot, precision recall (PR) plot, and gain curve indicates the co-ordinates when using the probability threshold that gives a sensitivity of 80% (probability threshold = 0·037). The positive predictive value (PPV) and negative predictive value (NPV) are also reported in the ROC plot using this threshold.

#### **Table S8.3** Coefficients of the **M6PD-A_0-6_ model**.

| **Variable** | **Coefficient** |
| --- | --- |
| Intercept | -2·753 |
| Weight for age z-score | -0·361 |
| MUAC | -0·219 |
| Time it took to reach hospital, >1 hour | 0·233 |
| Sucking well when breastfeeding, or feeding well if not breastfed | -0·206 |
| SpO_2_ | -0·149 |
| Duration of present illness, 8 days – 1 month | 0·130 |
| Duration of present illness, >1 month | 0·031 |
| Age | 0·031 |
| Age × Weight for age z-score | -0·064 |
| Age × Duration of present illness, 48 hours – 7 days | 0·005 |
| Age × Jaundice | 0·087 |

Interactions between variables are indicated by the multiplication sign.

Abbreviations: MUAC = mid-upper arm circumference; SpO_2_ = oxygen saturation

#### **Table S8.4** Coefficients of the **M6PD-A_6-60_ model**.

| **Variable** | **Coefficient** |
| --- | --- |
| Intercept | -3·257 |
| MUAC | -0·381 |
| Haemoglobin | -0·225 |
| Weight for age z-score | -0·187 |
| SpO_2_ | -0·176 |
| How long since last admission, <7 days | 0·072 |
| How long since last admission, 7 days – 1 month | 0·132 |
| How long since last admission, 1 month – 1 year | 0·020 |
| How long since last admission, >1 year | -0·058 |
| Water source, bore hole | 0·002 |
| Water source, municipal water | -0·087 |
| HIV+ | 0·102 |
| Age × How long since last admission, 7 days – 1 month | 0·003 |
| Age × How long since last admission, 1 month – 1 year | 0·067 |
| Age × Water source, bore hole | 0·164 |
| Age × Water source, municipal water | -0·036 |

Interactions between variables are indicated by the multiplication sign.

Abbreviations: HIV = human immunodeficiency virus; MUAC = mid-upper arm circumference; SpO_2_ = oxygen saturation

#### **Figure S8.3** Variable importance plot of the **M6PD-A_0-6_** model

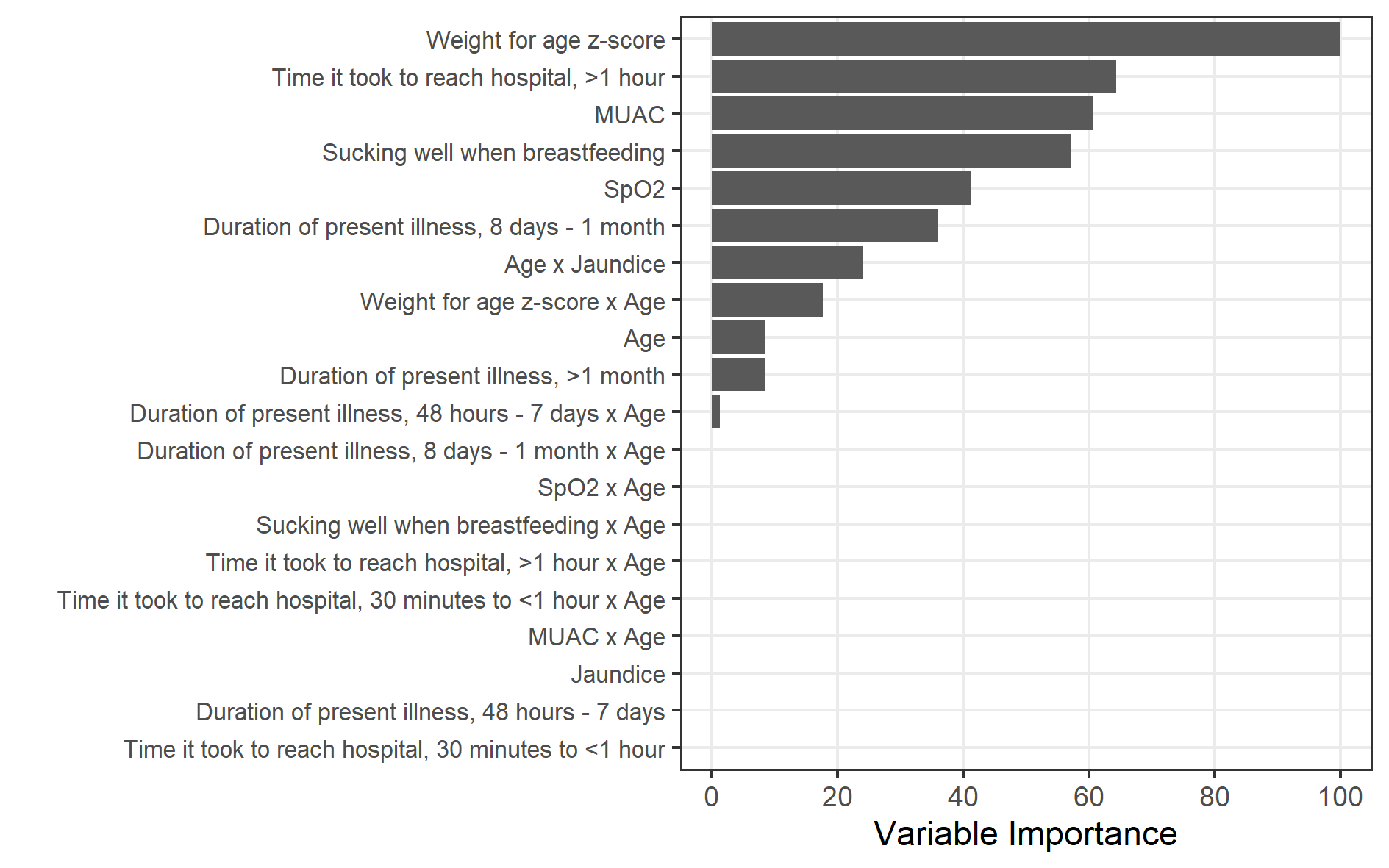

#### **Figure S8.4** Variable importance plot of the **M6PD-A_6-60_** model

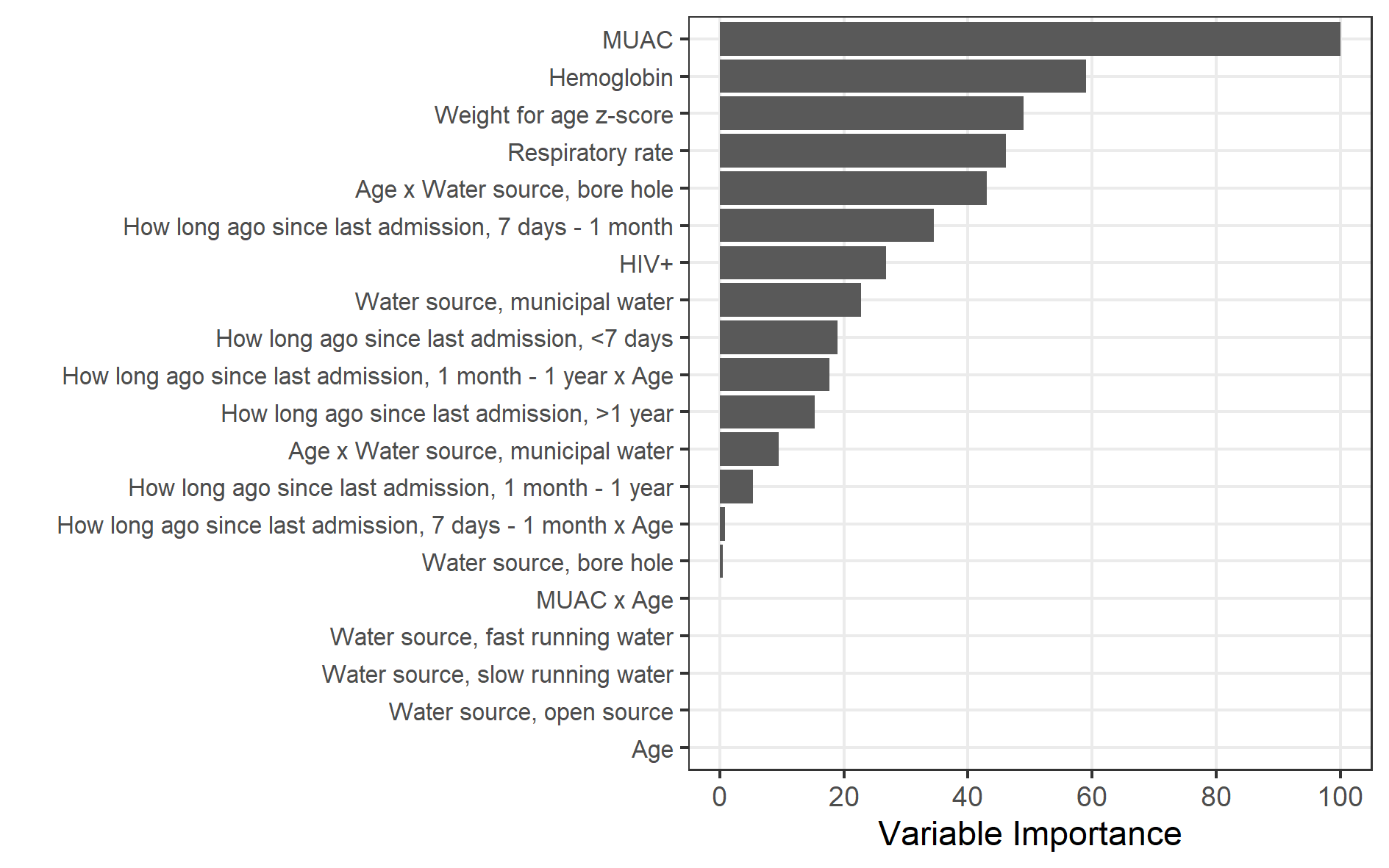
